# Immunogenicity and safety of NVX-CoV2373 as a homologous or heterologous booster: A phase 3 randomized clinical trial in adults

**DOI:** 10.1101/2023.03.16.23287030

**Authors:** Fritz Raiser, Matthew Davis, Jeffrey Adelglass, Miranda R. Cai, Gordon Chau, Shane Cloney-Clark, Mark Eickhoff, Raj Kalkeri, Irene McKnight, Joyce Plested, Mingzhu Zhu, Lisa Dunkle, 2019nCoV-307 study team

## Abstract

**Background:** To combat the SARS-CoV-2 pandemic, multiple vaccines using different manufacturing platforms have been developed, including NVX-CoV2373 (an adjuvanted recombinant protein vaccine). As SARS-CoV-2 variants have emerged, some of which evade vaccine-induced immunity, introduction of vaccine booster doses has become critical. Employing different vaccine types for primary series vaccination and boosting could expand vaccine coverage and access. This study assessed whether NVX-CoV2373 would induce similar responses when used as a heterologous or homologous booster.

**Methods:** The 2019nCoV-307 study was a phase 3, randomized, observer-blinded trial evaluating immunogenicity and safety of NVX-CoV2373 in previously vaccinated adults aged 18-49 years in the United States (NCT05463068). Participants were randomized 1:1:1 to receive one intramuscular injection of NVX-CoV2373 from one of three different manufacturing lots. Immunogenicity was assessed by immunoglobulin G (IgG) and neutralizing antibodies (NAb). These responses were compared for the three lots, and for participants with primary series with or without a prior booster dose of the mRNA-1273, BNT162b2, Ad26.COV2.S, or NVX- CoV2373 COVID-19 vaccines.

**Results:** A total of 911 participants were randomized between July 11 and 13, 2022, with 905 being assessed for safety and 848 for immunogenicity. Immunogenicity of NVX-CoV2373 met prespecified equivalence criteria between lots, and the booster dose was well-tolerated. NVX- CoV2373 induced robust IgG and NAb responses when used as a first or later booster dose, regardless of primary series vaccine type. Seroconversion rates were also similar across previous vaccine types. Induced antibodies were strongly reactive, even to the immune-evasive Omicron BA.1 and BA.5 variants.

**Conclusions:** NVX-CoV2373 showed consistent immunogenicity between lots, with no new safety signals identified. Use of NVX-CoV2373 as a homologous or heterologous booster dose (first or later) is supported.

## 1. Introduction

The unprecedented response to the SARS-CoV-2 pandemic, involving a global public-private- nonprofit collaboration, resulted in world-wide availability of vaccines based on multiple manufacturing platforms within less than 2 years after identification of the pandemic pathogen [1–3]. Following initial widespread vaccine availability, the SARS-CoV-2 virus began evolving through escape mutations that resulted in further broadening of transmission and breakthrough infections [4–7]. The need for widespread access to additional vaccine doses to be used as boosters led to the assessment of effectiveness of employing several different vaccine formats to ensure broad coverage and offer choice of vaccine format for the populace [8, 9].

The adjuvanted recombinant spike protein (rS) vaccine NVX-CoV2373 (Novavax SARS-CoV-2 rS, adjuvanted) demonstrated approximately 90% efficacy against COVID-19 in a phase 3 study of almost 30,000 adults [10]. The 2019nCoV-307 study (Clinicaltrials.gov: NCT05463068) was intended to demonstrate the consistency of immunogenicity induced by multiple manufacturing lots of NVX-CoV2373. In addition, the study assessed immunogenicity induced by a single injection of NVX-CoV2373 following prior priming with or without boosting by a U.S. Food and Drug Administration (FDA)-authorized or approved vaccine. This type of lot consistency study design is required for full approval of a vaccine in the United States (US); however, it later became apparent that the immune responses to NVX-CoV2373 used as a homologous or heterologous booster were of significant public health interest, and thus these results will be the focus of this manuscript.

We report the consistent and robust immune response induced by multiple manufacturing lots of NVX-CoV2373 when administered to US participants as a first or later booster following initial priming with NVX-CoV2373 (homologous boost) or other authorized vaccines (heterologous boost). The majority of the study results reflected heterologous boosting following earlier vaccination with authorized or approved mRNA vaccines.

## 2. Methods

### 2.1 Study design, participants, oversight

This study (2019nCoV-307) was a phase 3, observer-blinded, randomized trial in the US comparing the immunogenicity and safety of three different lots of NVX-CoV2373 (Novavax, Gaithersburg, MD) in previously vaccinated adults aged 18 to 49 years. Participants from 31 study centers in the US were enrolled and randomized between July 11 and 13, 2022. Eligible participants were medically stable adults aged 18 to 49 years who had been previously vaccinated against SARS-CoV-2 with at least a full primary series and optionally, a booster dose. The most recent dose was required to have been received at least 6 months prior to study vaccination. Key exclusion criteria included: pregnancy/lactation, autoimmune or immunodeficiency disease/condition/therapy, history of COVID-19 infection ≤4 months prior to randomization, immunosuppressed or immunocompromised status, allergy/anaphylaxis to vaccine components, history of myocarditis/pericarditis, and receipt of any vaccine 90 days or less prior to study vaccination (except influenza or rabies vaccines).

Enrolled participants had previously received one of the following: two or three doses of NVX- CoV2373, a full course of an FDA-authorized/approved COVID-19 vaccine with or without a booster dose (the mRNA vaccines mRNA-1273 [Moderna, Cambridge, MA] or BNT162b2 [Pfizer, New York, NY], or the adenoviral vector vaccine Ad26.COV2.S [Janssen Pharmaceuticals, Beerse, Belgium]), or a full course of these COVID-19 vaccines with or without a heterologous booster dose. In this study, participants were randomized in a 1:1:1 ratio to receive one 0.5 mL intramuscular injection of NVX-CoV2373 (5 ug SARS-CoV-2 rS + 50 ug Matrix-M™ adjuvant) in the deltoid muscle from one of three different manufacturing lots (Novavax Lot/PCI WO Number: Group 1 - 4302MF003/1908368, Group 2 - 4302MF010/1908370, Group 3 - 4302MF013/1908372) using a 1” 25-gauge needle. Participants were followed through 28 days after the vaccination for immunogenicity and safety outcomes.

Both clinical study personnel and participants were blinded to the lot assignment (see Supplement). Unblinded study pharmacy personnel managed vaccine logistics, preparation, and administration, but were not involved in study-related assessments or participant contact for data collection. Novavax was the trial sponsor and was responsible for study design and development. Clinical trial vaccine antigen was manufactured by Serum Institute of India and the adjuvanted vaccine material was formulated and packaged by PCI Pharma Services (Philadelphia, PA) through partnerships with Novavax. Additional details of trial design, conduct, oversight, and analyses are available at: https://clinicaltrials.gov/ct2/show/NCT05463068. The protocol and amendments were approved and overseen by a central institutional review board. Participants provided written informed consent before entering the study, and the study was conducted in compliance with the Declaration of Helsinki, the International Conference on Harmonization Guidelines for Good Clinical Practice, and applicable national and local regulations. Study team members are listed in **Table S1**.

### 2.2 Immunogenicity assessments

The primary immunogenicity endpoint was to demonstrate equivalence of immunoglobulin G (IgG) responses to SARS-CoV-2 rS-protein at Day 29 for the three different manufacturing lots, as measured by geometric mean enzyme-linked immunosorbent assay (ELISA) units (GMEU). Equivalence was met if the 95% confidence intervals (CIs) of the GMEU ratios (GMEUR) for the pairwise comparisons of all three lots were between 0.67 and 1.5. A key secondary endpoint was to determine the seroconversion rate (SCR), defined as the proportion of participants who showed ≥ 4-fold increases from baseline in anti-rS IgG concentrations. An ad hoc analysis assessed differences in IgG responses in participants according to the type of prior primary series received and booster history. Validated anti-rS IgG ELISA assays were performed as previously published [11]; 95% CIs were calculated by exponentiating the log-transformed 95% CI of the mean (for GMEUs), paired t-distribution (for GMFRs), Clopper-Pearson methods (SCRs), or Miettinen and Nurminen method (SCR differences).

Neutralizing antibody (NAb) responses to SARS-CoV-2 S-protein at Day 29 were also assessed, using geometric mean titers (GMTs) in a microneutralization assay and by SCR, as for IgG responses. Validated live virus-based microneutralization assays (MN_50_) assays were performed as previously published [12]. 95% CIs were calculated by exponentiating the log-transformed 95% CI of the mean (for GMTs), paired t-distribution (for GMFRs), Clopper-Pearson methods (SCRs), or Miettinen and Nurminen method (SCR differences).

The immunogenicity endpoints were assessed in the Per-Protocol (PP) Analysis Set, which included all participants who received the study vaccine according to the protocol, had serology results for Day 1 and Day 29 (Day 0 and Day 28 post-vaccination, respectively), and without any major protocol violations that could impact immune responses.

### 2.3 Safety Assessments

Safety outcomes were assessed in the Safety Analysis Set, which included all participants who provided consent, were randomized/enrolled, and received one dose of study vaccine. Safety Analysis Set participants were analyzed based on actual treatment received. Outcomes assessed included unsolicited adverse events (AEs), AEs of special interest (AESIs), and medically attended AEs (MAAEs) through Day 29; serious AEs (SAEs), AEs leading to discontinuation, and deaths through the end of the study. AESIs included potentially immune-mediated medical conditions, myocarditis/pericarditis, and AEs related to COVID-19.

## 3. Results

### 3.1 Participants

Between July 11 and 13, 2022, a total of 990 participants were screened for eligibility, and 911 were enrolled and randomized (**Fig. 1**). The Safety Analysis Set included 905 participants (n=298 for Group 1, n=303 for Group 2, n=304 for Group 3). The PP Analysis Set included 848 participants (n=275 for Lot 1, n=283 for Lot 2, n=290 for Lot 3). For the PP Analysis Set, mean (SD) age was 36.6 (8.41) years, 58.4% were female, 74.5% were White, and 4.8% had documented previous SARS-CoV-2 infection. Most participants had previously received an mRNA vaccine primary series (mRNA-1273 [34.1%] or BNT162b2 [57.4%]) (**Table 1**). Baseline demographics in the Safety Analysis Set were also well-balanced across treatment groups (**Table S2**).

**Table 1.** Demographics and baseline characteristics (Per Protocol Analysis Set^a^).

|  | **Group 1**<br>**N=275**<br>**n (%)** | **Group 2**<br>**N=283**<br>**n (%)** | **Group 3**<br>**N=290**<br>**n (%)** | **Total**<br>**N=848**<br>**n (%)** |
| --- | --- | --- | --- | --- |
| **Age (years)<sup>b</sup>** |  |  |  |  |
| n | 275 | 283 | 290 | 848 |
| Mean (SD) | 36.3 (8.29) | 36.8 (8.80) | 36.6 (8.15) | 36.6 (8.41) |
| Median | 38.0 | 38.0 | 38.0 | 38.0 |
| Min, Max | (19, 49) | (18, 49) | (18, 49) | (18, 49) |
| **Sex, n (%)** |  |  |  |  |
| Male | 114 (41.5) | 127 (44.9) | 112 (38.6) | 353 (41.6) |
| Female | 161 (58.5) | 156 (55.1) | 178 (61.4) | 495 (58.4) |
| **Ethnicity, n (%)** |  |  |  |  |
| Hispanic or Latino | 44 (16.0) | 39 (13.8) | 49 (16.9) | 132 (15.6) |
| Not Hispanic or Latino | 228 (82.9) | 241 (85.2) | 239 (82.4) | 708 (83.5) |
| Not Reported/ Specified | 3 (1.1) | 3 (1.1) | 2 (0.7) | 8 (0.9) |
| **Race, n (%)** |  |  |  |  |
| American Indian or Alaska Native | 3 (1.1) | 1 (0.4) | 3 (1.0) | 7 (0.8) |
| Asian | 9 (3.3) | 10 (3.5) | 9 (3.1) | 28 (3.3) |
| Black or African American | 49 (17.8) | 55 (19.4) | 48 (16.6) | 152 (17.9) |
| Native Hawaiian or Other Pacific Islander | 0 | 0 | 3 (1.0) | 3 (0.4) |
| White | 205 (74.5) | 209 (73.9) | 218 (75.2) | 632 (74.5) |
| Other | 6 (2.2) | 3 (1.1) | 8 (2.8) | 17 (2.0) |
| Not Reported/ Specified | 3 (1.1) | 5 (1.8) | 1 (0.3) | 9 (1.1) |
| **BMI, n (%)** |  |  |  |  |
| Underweight (<18.5) | 4 (1.5) | 4 (1.4) | 4 (1.4) | 12 (1.4) |
| Healthy (18.5-24.9) | 54 (19.6) | 72 (25.4) | 68 (23.4) | 194 (22.9) |
| Overweight (25.0-29.9) | 75 (27.3) | 81 (28.6) | 62 (21.4) | 218 (25.7) |
| Obese (≥30.0) | 142 (51.6) | 126 (44.5) | 156 (53.8) | 424 (50.0) |
| **Time between first dose of previous COVID-19 vaccine and Day 1 dose (weeks)** |  |  |  |  |
| n | 275 | 283 | 290 | 848 |
| Mean (SD) | 66.6 (13.35) | 68.5 (13.98) | 68.1 (11.96) | 67.7 (13.12) |

|  | **Group 1**<br>**N=275**<br>**n (%)** | **Group 2**<br>**N=283**<br>**n (%)** | **Group 3**<br>**N=290**<br>**n (%)** | **Total**<br>**N=848**<br>**n (%)** |
| --- | --- | --- | --- | --- |
| Median | 67.7 | 67.9 | 68.4 | 68.1 |
| Min, Max | (26, 102) | (29, 101) | (29, 102) | (26, 102) |
| **Previous SARS-CoV-2 infection, n (%)** |  |  |  |  |
| Yes | 17 (6.2) | 12 (4.2) | 12 (4.1) | 41 (4.8) |
| **Baseline PCR, n(%)** |  |  |  |  |
| Positive | 7 (2.5) | 8 (2.8) | 6 (2.1) | 21 (2.5) |
| Negative | 261 (94.9) | 268 (94.7) | 276 (95.2) | 805 (94.9) |
| Missing | 7 (2.5) | 7 (2.5) | 8 (2.8) | 22 (2.6) |
| **Regimen of previous COVID-19 vaccines received before the study, n (%)** |  |  |  |  |
| **NVX-CoV2373** |  |  |  |  |
| Primary vaccination series (no booster) | 1 (0.4) | 4 (1.4) | 2 (0.7) | 7 (0.8) |
| Primary series + NVX-CoV2373 booster (homologous boost) | 1 (0.4) | 2 (0.7) | 1 (0.3) | 4 (0.5) |
| Primary series + other booster (heterologous boost) | 4 (1.5) | 4 (1.4) | 4 (1.4) | 12 (1.4) |
| **mRNA-1273** |  |  |  |  |
| Primary vaccination series (no booster) | 42 (15.3) | 42 (14.8) | 47 (16.2) | 131 (15.4) |
| Primary series + mRNA-1273 booster (homologous boost) | 50 (18.2) | 43 (15.2) | 41 (14.1) | 134 (15.8) |
| Primary series + other booster (heterologous boost) | 6 (2.2) | 7 (2.5) | 11 (3.8) | 24 (2.8) |
| **BNT162b2** |  |  |  |  |
| Primary vaccination series (no booster) | 65 (23.6) | 67 (23.7) | 58 (20.0) | 190 (22.4) |
| Primary series + BNT162b2 booster (homologous boost) | 74 (26.9) | 85 (30.0) | 101 (34.8) | 260 (30.7) |
| Primary series + other booster (heterologous boost) | 10 (3.6) | 12 (4.2) | 15 (5.2) | 37 (4.4) |
| **Ad26.COV2.S** |  |  |  |  |
| Primary vaccination series (no booster) | 7 (2.5) | 6 (2.1) | 6 (2.1) | 19 (2.2) |
| Primary series + Ad26.COV2.S booster (homologous boost) | 4 (1.5) | 1 (0.4) | 1 (0.3) | 6 (0.7) |
| Primary series + other booster (heterologous boost) | 7 (2.5) | 8 (2.8) | 3 (1.0) | 18 (2.1) |
| **Other** | 4 (1.5) | 2 (0.7) | 0 | 6 (0.7) |

BMI, body mass index; PCR, polymerase chain reaction; SD, standard deviation.
<sup>a</sup>Per Protocol Analysis Set includes all participants who receive the study vaccine according to the protocol, have serology results for Day 1 and Day 29 available after the vaccination, and have no major protocol violations. Treatment group in Per Protocol Analysis Set is based on the actual dose(s) received. N is the number of participants in Per Protocol Analysis Set; n is the number of participants in each specified category.
<sup>b</sup>Age is calculated as the lowest integer result of (Date of Study Day 1 – Date of Birth + 1)/365.25.

**Fig. 1.**
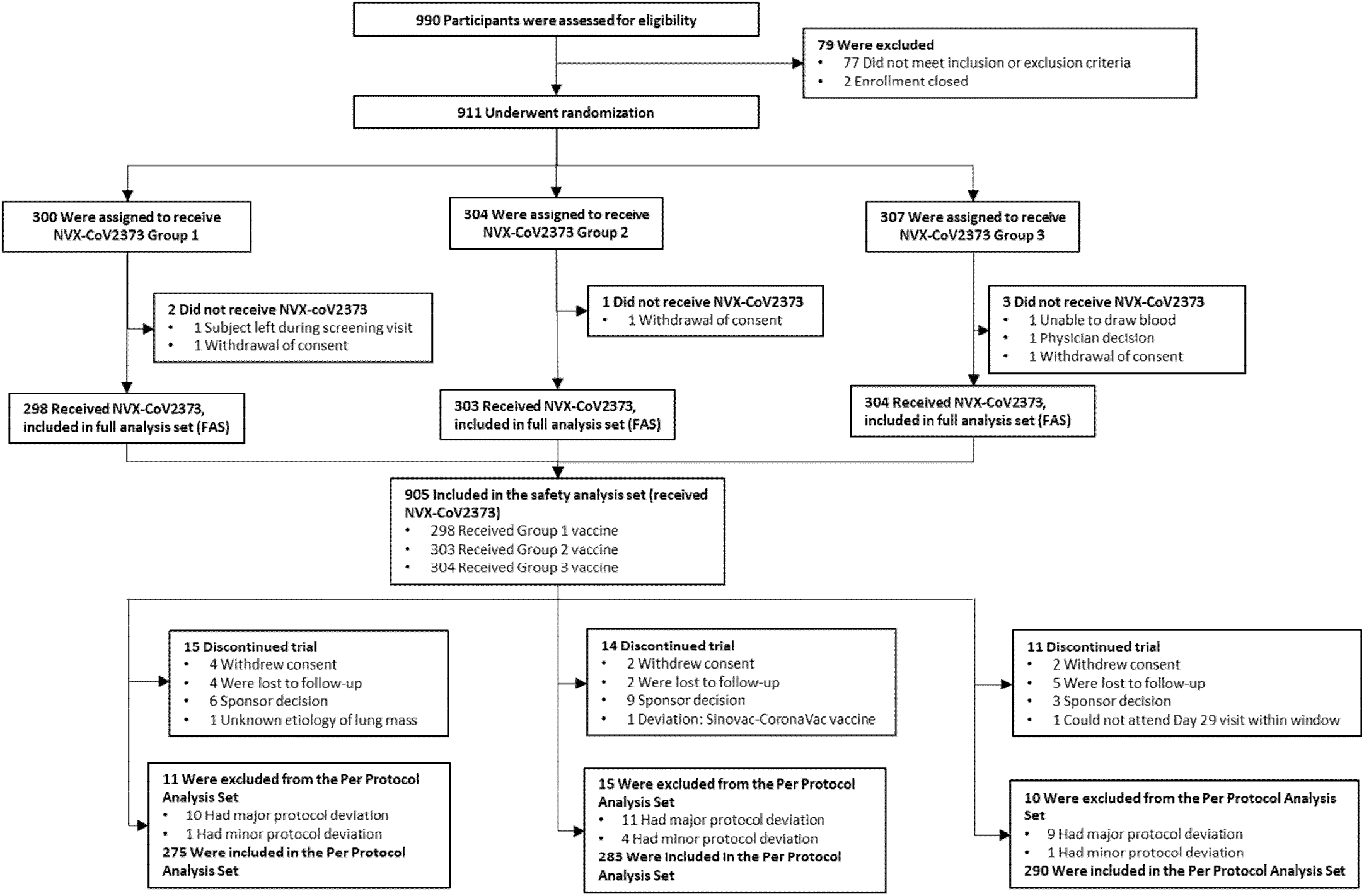
CONSORT diagram. Participant flow diagram for the trial.

### 3.2 NVX-CoV2373 lot-to-lot consistency

NVX-CoV2373 induced equivalent IgG immune responses across the three different commercially manufactured lots, meeting the primary endpoint of the study. IgG GMEUs (95% CIs) ranged from 33233.4 (28863.0–38265.4) to 36142.6 (31430.8–41560.8). Pairwise comparisons of IgG GMEURs were within the predefined equivalence criteria (range of 0.67 to 1.5). SCRs were similar between lots for IgG responses (**Figs. S1A-B, Table S3**). GMFRs and SCRs were also similar between lots for NAb responses (**Fig. S1C-D, Table S4**), meeting the secondary endpoints.

### 3.3 Immunogenicity by Prior Vaccine History

IgG and NAb responses to SARS-CoV-2 rS-protein following a booster dose of NVX-CoV2373 were assessed based on type of primary series received before the study, and whether a booster dose had been received before the study.

#### 3.3.1 No prior booster history

A total of 347 participants had received primary series but no prior booster doses before the study (n=190 for BNT162b2, n=131 for mRNA-1273, n=19 for Ad26.COV2.S, n=7 for NVX- CoV2373) (**Table 1**). At Day 1 (prior to receiving the NVX-CoV2373 dose), the participants in the NVX-CoV2373 group had generally higher anti-S IgG GMEUs (although the 95% CI was wide) (**Fig. 2A, Table S5**). At Day 29 (after receiving the study vaccine NVX-CoV2373 as one booster dose), participants in all vaccine type groups showed similar increases in IgG levels. The participants who had received an NVX-CoV2373 primary series before the study (and thus received the study vaccine as a homologous booster) showed the greatest fold-rise from pre-booster dose to post-booster dose, although the sample size of this group was small and this group had higher baseline levels than other groups.

**Fig. 2.**
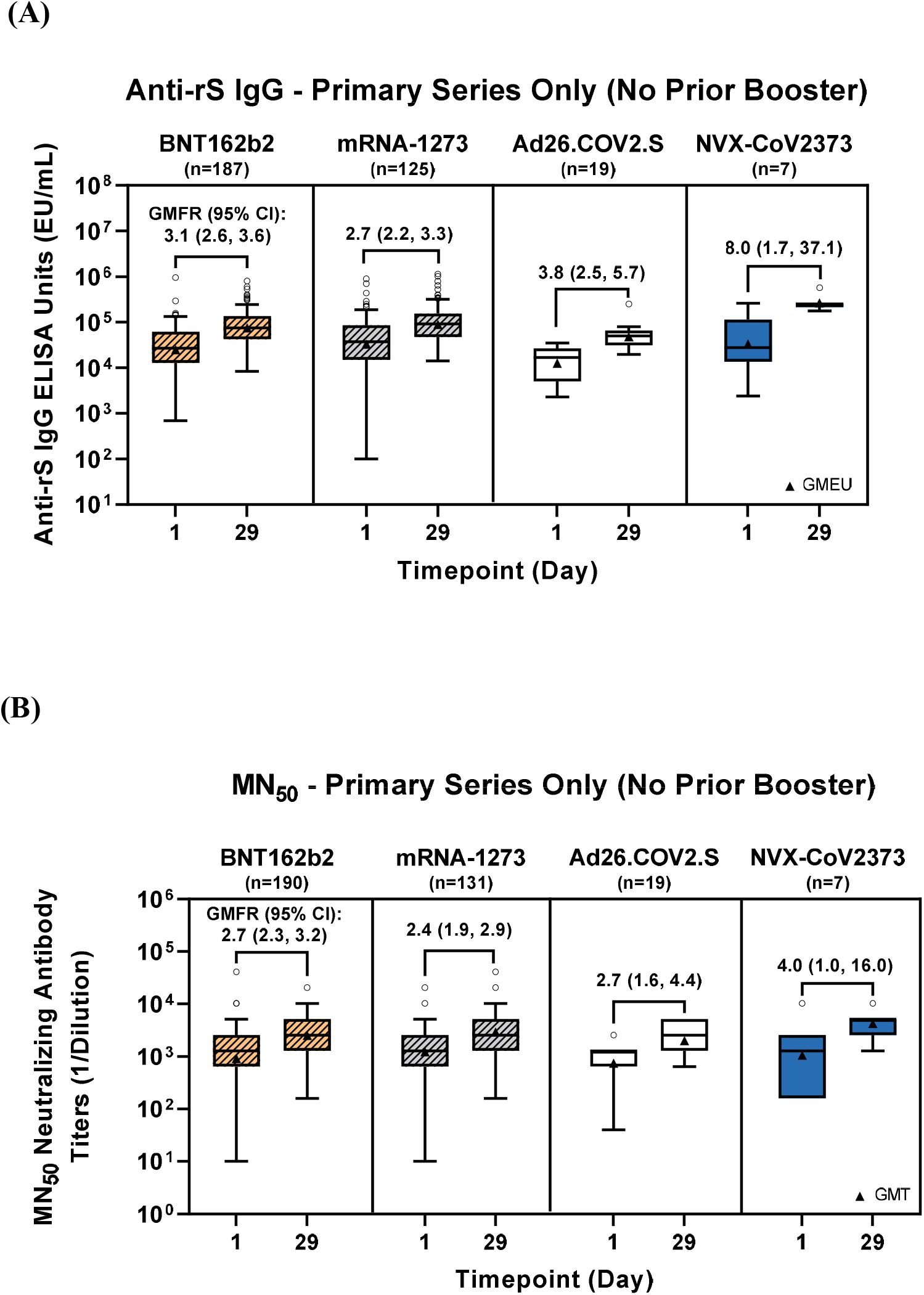
Anti-rS IgG and neutralizing antibody (MN) responses before and after boosting with NVX-CoV2373 > 6 months after primary series (participants with prior primary series only, no prior booster). Immunogenicity of NVX-CoV2373 in participants with only a prior primary series (no booster) was assessed by (A) Anti-rS IgG responses measured by a validated ELISA assay, and (B) neutralizing antibody responses measured by a validated live virus-based microneutralization assay. ELISA units or titers are graphed as boxplots with GMEU/GMT as triangles (whiskers drawn using the Tukey method, values outside the whiskers are shown as open circles), and GMFRs with 95% CIs are shown above the boxes. CI, confidence interval; ELISA, enzyme-linked immunosorbent assay; GMEU, geometric mean ELISA units; GMFR, geometric mean fold-rise; GMT, geometric mean titer; IgG, immunoglobulin G; IQR, interquartile range; MN, microneutralization.

NAb responses at Day 1 were at similar levels for participants in each group. The NVX- CoV2373 booster increased NAbs at Day 29 regardless of prior primary series type. GMFRs showed similar increases for all vaccine type groups; those in the NVX-CoV2373 primary series group appeared to have the greatest fold-rise in response to the NVX-CoV2373 boost (**Fig. 2B, Table S6**).

#### 3.3.2 Prior booster history

Of those participants who had previously been boosted (n=495), the majority had received an mRNA primary series (BNT162b2 or mRNA-1273) and had received a homologous booster prior to participation in this study (n=394). Those who received Ad26.COV2.S or NVX- CoV2373 more frequently had received a heterologous booster (n=30) prior to this study (**Table 1**). At Day 1, participants in the two homologous prior booster groups had similar levels of anti-rS IgG (**Table S7**), whereas participants with a heterologous prior booster after an NVX- CoV2373 primary series had higher IgG GMEUs at baseline of this study(**Table S8**). At Day 29, all vaccine type groups showed similar increases in IgG levels (**Fig. 3A-B, Table S7-8**). Among those who had received a prior homologous booster (same primary series and booster type), those receiving earlier doses of NVX-CoV2373 showed the highest GMEUs in response to the NVX-CoV2373 dose in this study, despite baseline levels similar to other groups (**Fig. 3A, Table S7**). Among those who had previously received a primary series followed by a heterologous booster, participants who previously had an NVX-CoV2373 primary series and another booster had the highest IgG levels, which may be related to the higher baseline levels (**Fig. 3B, Table S8**).

**Fig. 3.**
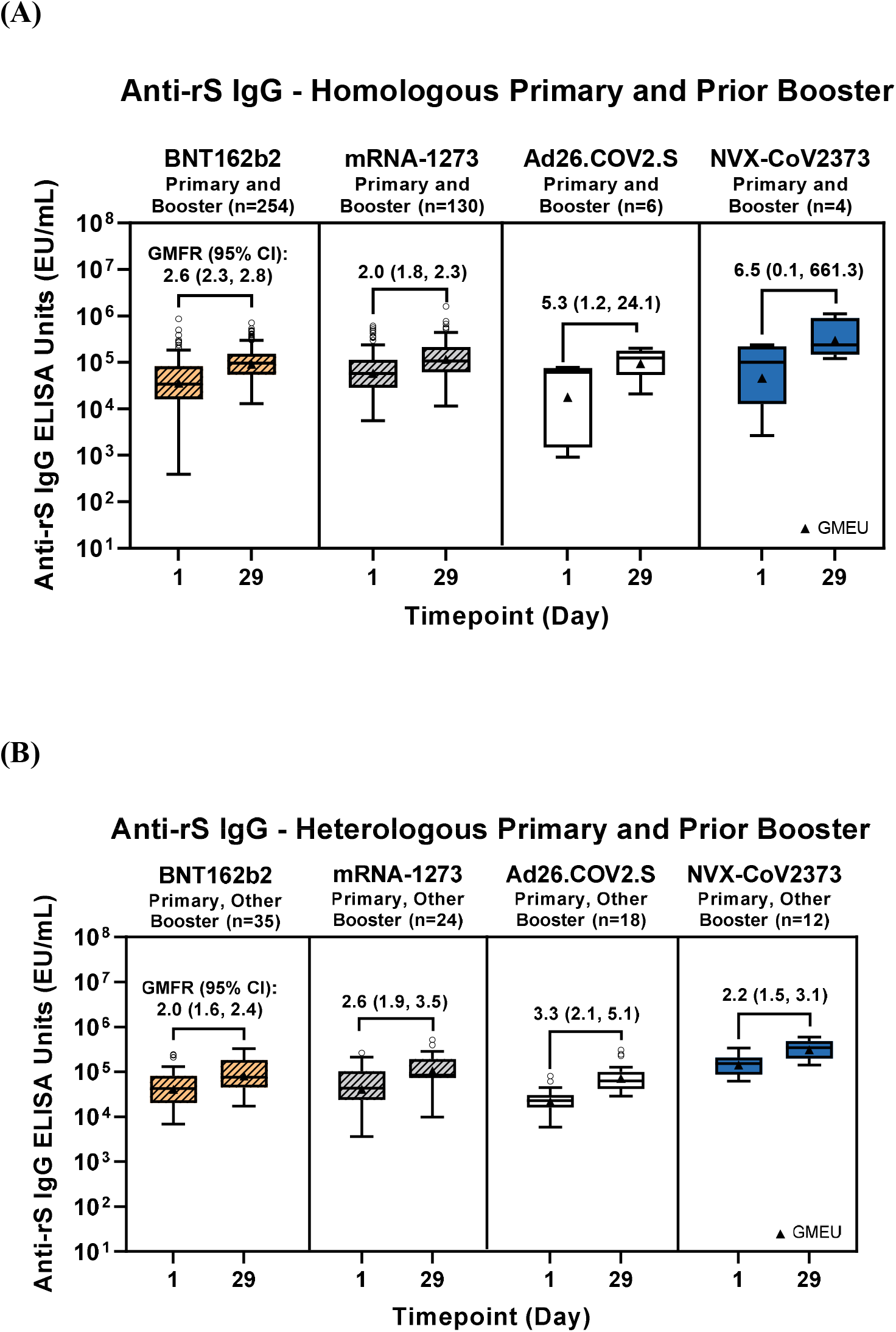
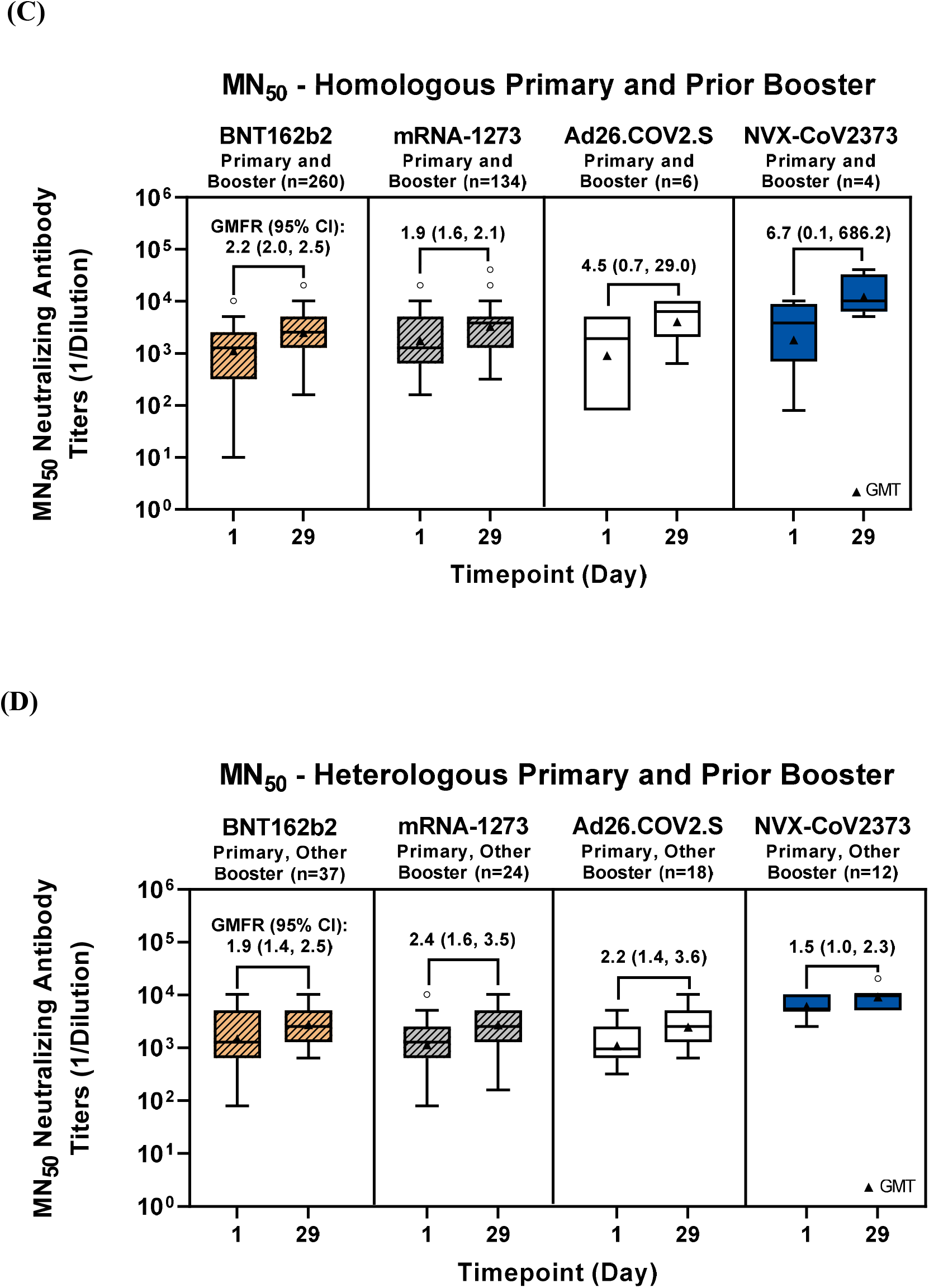
Anti-rS IgG and MN responses before and after boosting with NVX-CoV2373 (participants with primary series and prior homologous or heterologous boosting). Immunogenicity of NVX-CoV2373 in participants with a prior primary series and a homologous or heterologous prior booster was assessed by (A-B) Anti-rS IgG responses measured by a validated ELISA assay, and (C-D) neutralizing antibody responses measured by a validated live virus-based microneutralization assay. ELISA units or titers are graphed as boxplots with GMEU/GMT as triangles (whiskers drawn using the Tukey method, values outside the whiskers are shown as open circles), and GMFRs with 95% CIs are shown above the boxes. CI, confidence interval; ELISA, enzyme-linked immunosorbent assay; GMEU, geometric mean ELISA units; GMFR, geometric mean fold-rise; GMT, geometric mean titer; IgG, immunoglobulin G; IQR, interquartile range; MN, microneutralization.

NAb levels were similar between groups at Day 1, and showed similar levels across groups at Day 29 after receipt of the NVX-CoV2373 dose in this study (**Fig. 3C, Table S9**). The relatively small number of participants who had previously received a primary series of NVX-CoV2373 and a homologous booster showed the highest response to the NVX-CoV2373 dose in this study, which may be due in part to having somewhat higher baseline levels than some of the other groups. The increase in NAbs was similar between all vaccine type groups for those with a previous primary series and homologous booster. Among those who had previously received a primary series and heterologous booster, the increase in NAbs in this study was similar regardless of prior vaccine types (**Fig. 3D, Table S10**).

#### 3.3.3. Seroconversion rates (SCR) after NVX-CoV2373 booster

SCRs after the dose of NVX-CoV2373 in this study (defined as the proportion of participants with ≥ 4-fold increases from baseline anti-rS IgG or NAb levels) were also assessed. For IgG responses, SCRs were generally similar (with overlapping 95% CIs) for each vaccine type group within their respective boosting history groups. Participants who previously had received a heterologous booster generally exhibited lower SCRs than participants who had received a prior homologous booster or no prior booster (**Fig. 4A, Tables S5, S7–8**). NAb SCRs were similar among the vaccine groups within each booster category. While SCRs were similar for recipients of prior mRNA vaccines regardless of booster history (approximately 35% to 36% for BNT162b2 and 27% to 33% for mRNA-1273), there was greater variability in SCRs for the non-mRNA vaccines (**Fig. 4B, Table S6, S9-10**).

**Figure 4.**
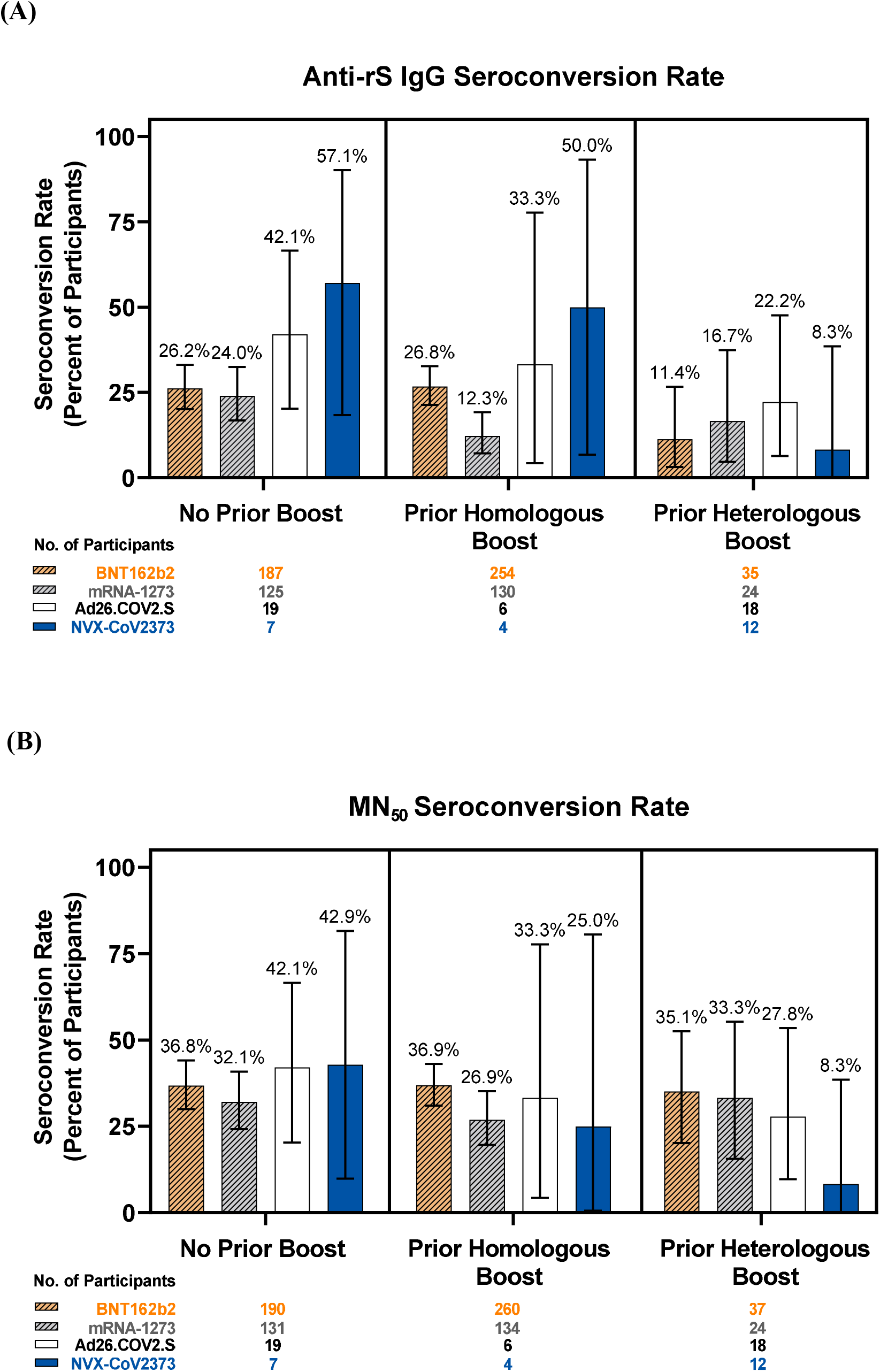
SARS-CoV-2 import risk into San Diego. **(A)** Weekly estimated number of travelers arriving into San Diego from January 2020-June 2021. Locations are sorted by the total number of estimated visitors over this period and only the top 25 are shown. Location names are styled depending on their administrative level: California counties are italicized, countries are bolded, and US states weighted normally (**B**) Scatter plot of each locations’ total estimated travelers into San Diego and the relative standard deviation in estimated travelers for the period indicated by panel A. The five locations with the greatest total estimated travelers into San Diego are highlighted in blue. (**C**) Proportion of travelers arriving into San Diego from the five locations with the greatest total estimated travelers (top-most five locations in panel A). (**D**) import risk into San Diego. import risk was estimated based on the number of infectious travelers relative to the population size and the total number of travelers at the origin. Only the five locations with the greatest total import risk into San Diego are shown. All other locations are colored in gray. (**E**) Relative import risk into San Diego. Locations are colored as in panel D, with gray representing all locations outside the top five locations.

#### 3.3.4 Antibody responses cross-reactive with viral variants after NVX-CoV2373 booster

IgG responses cross-reactive to the Omicron BA.1 and BA.5 variants were examined following receipt of the NVX-CoV2373 booster in this study for participants who had previously received an mRNA (mRNA-1273 or BNT162b2) or NVX-CoV2373 primary series alone. Levels of IgG were lower for participants in all vaccine type groups in response to Omicron BA.1/BA.5 variants than for the ancestral strain (**Fig. 5A, Table S11**). NVX-CoV2373 recipients showed higher GMEUs following the NVX-CoV2373 dose for the ancestral strain, the Omicron BA.1 strain, and the Omicron BA.5 strain compared to the mRNA vaccine recipients, which may be related to somewhat higher baseline IgG levels. The small number of NVX-CoV2373 primary series recipients showed the highest GMFRs for all variants . SCRs were generally similar between the mRNA vaccine types across IgG responses for different strains: ancestral (28.1%, 37.5%), Omicron BA.1 (37.5%, 45.0%), and Omicron BA.5 (37.5%, 47.5%) (**Fig. 5B, Table S11**). Recipients of NVX-CoV2373 primary series showed the highest SCRs, at 71.4% for the Omicron variants and 57.1% for the ancestral strain (although 95% CIs were wide and overlapping). This was due to NVX-CoV2373 primary series recipients reaching the highest IgG levels after the booster dose.

**Fig. 5.**
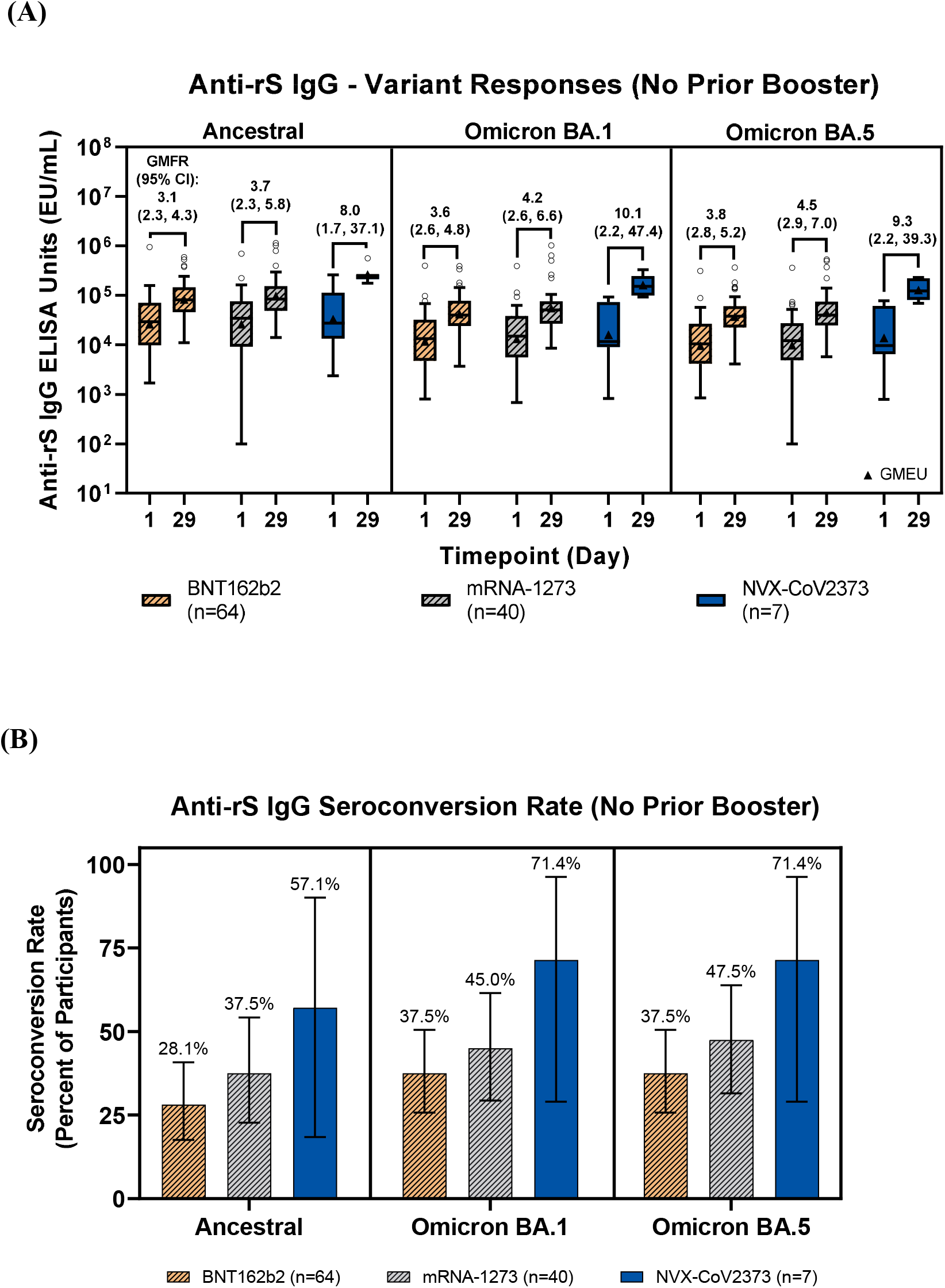
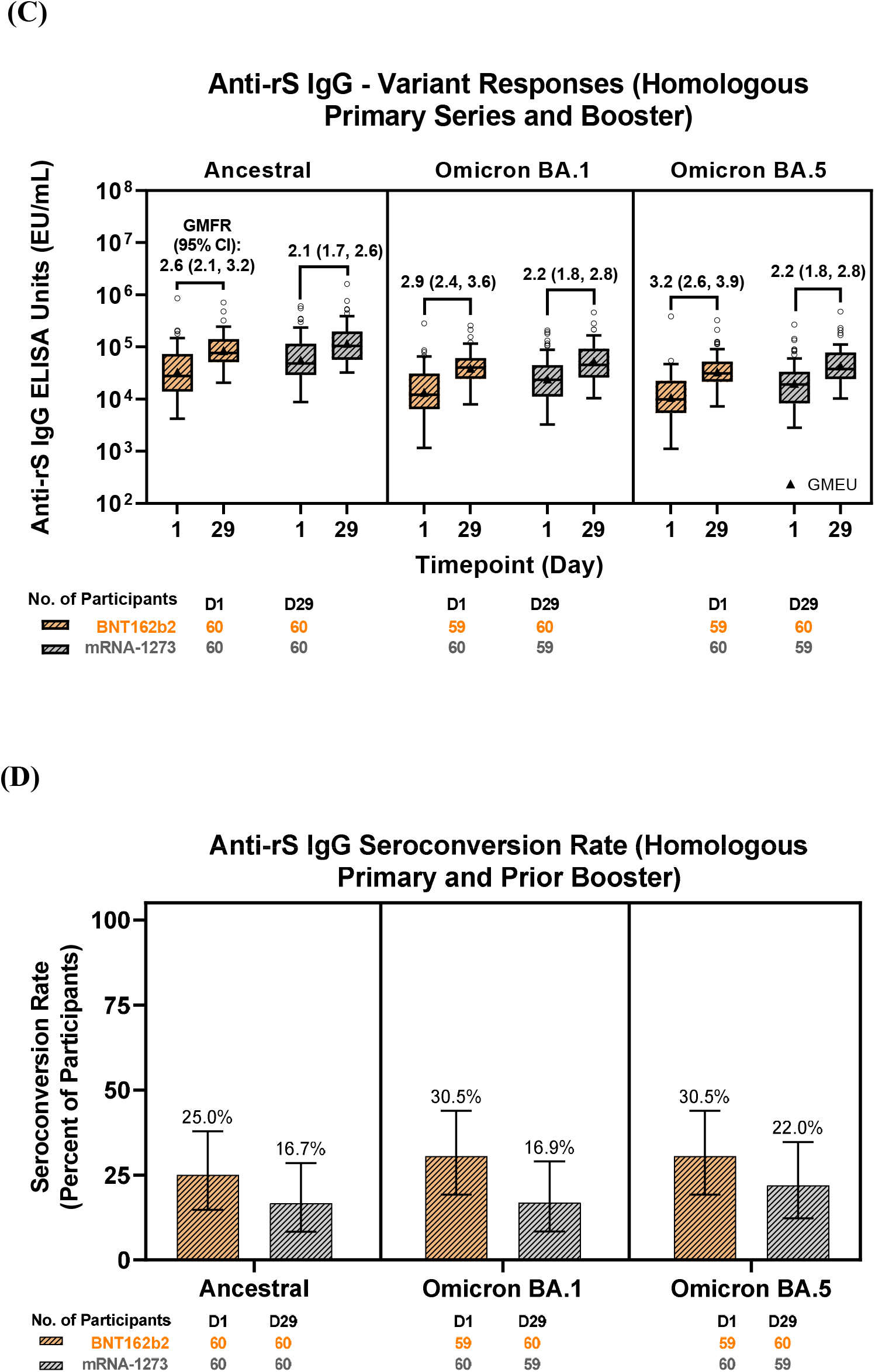
Anti-rS IgG responses to SARS-CoV-2 ancestral strain or variants after boosting with NVX-CoV2373. Immunogenicity of NVX-CoV2373 based on IgG responses to Ancestral strain or Omicron variants (BA.1, BA.5) was assessed. (A) Anti-rS IgG responses and (B) seroconversion rates for participants with only a prior primary series (no booster), and (C-D) for participants with a primary series and prior homologous booster, are shown. For anti-rS IgG responses, ELISA units are graphed as boxplots with GMEU as triangles (whiskers drawn using the Tukey method, values outside the whiskers are shown as open circles), and GMFRs with 95% CIs are shown above the boxes. For seroconversion, percentages are graphed with 95% CI. CI, confidence interval; ELISA, enzyme-linked immunosorbent assay; GMEU, geometric mean ELISA units; GMFR, geometric mean fold-rise; IgG, immunoglobulin G; IQR, interquartile range.

Immune responses for different viral strains (ancestral, Omicron BA.1, and Omicron BA.5) were also assessed in participants who had previously received an mRNA primary series and homologous booster. IgG levels increased following the receipt of NVX-CoV2373 in this study, with similar post-vaccination GMEUs for each variant across the recipients of the two prior mRNA vaccine types(**Fig. 5C, Table S12**). Among those in the BNT162b2 group and the mRNA-1273 group, SCRs following the NVX-CoV2373 dose in this study were similar across assays of different ancestral or variant strains (BNT: 16.7%, mRNA: 25.0%), Omicron BA.1 (16.9%, 30.5%), and Omicron BA.5 (22.0%, 30.5%) (**Fig. 5D, Table S12**).

### 3.4 Safety

A total of 39 (4.3%) participants reported unsolicited AEs. Only 6 (0.7%) were considered related to NVX-CoV2373, all of which were consistent with symptoms of reactogenicity. Two (0.2%) were severe, and 2 (0.2%) met regulatory criteria for serious AEs; none of these 4 events was considered related to study vaccine. No deaths, AESIs, treatment-related SAEs/severe AEs, or AEs leading to discontinuation were reported (**Table S13**). AEs were also collected by previous vaccine type (**Table 2**).

**Table 2.**
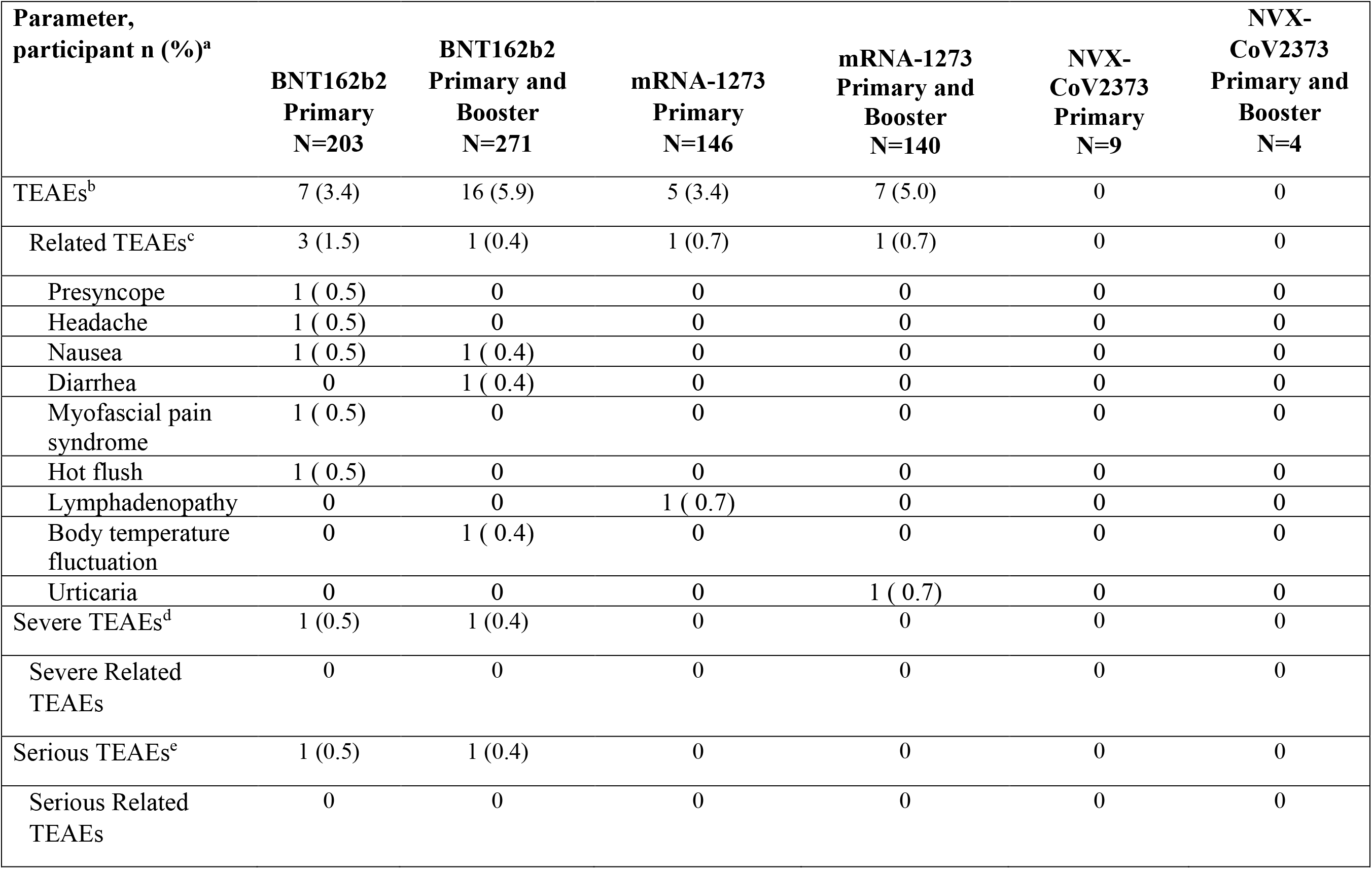
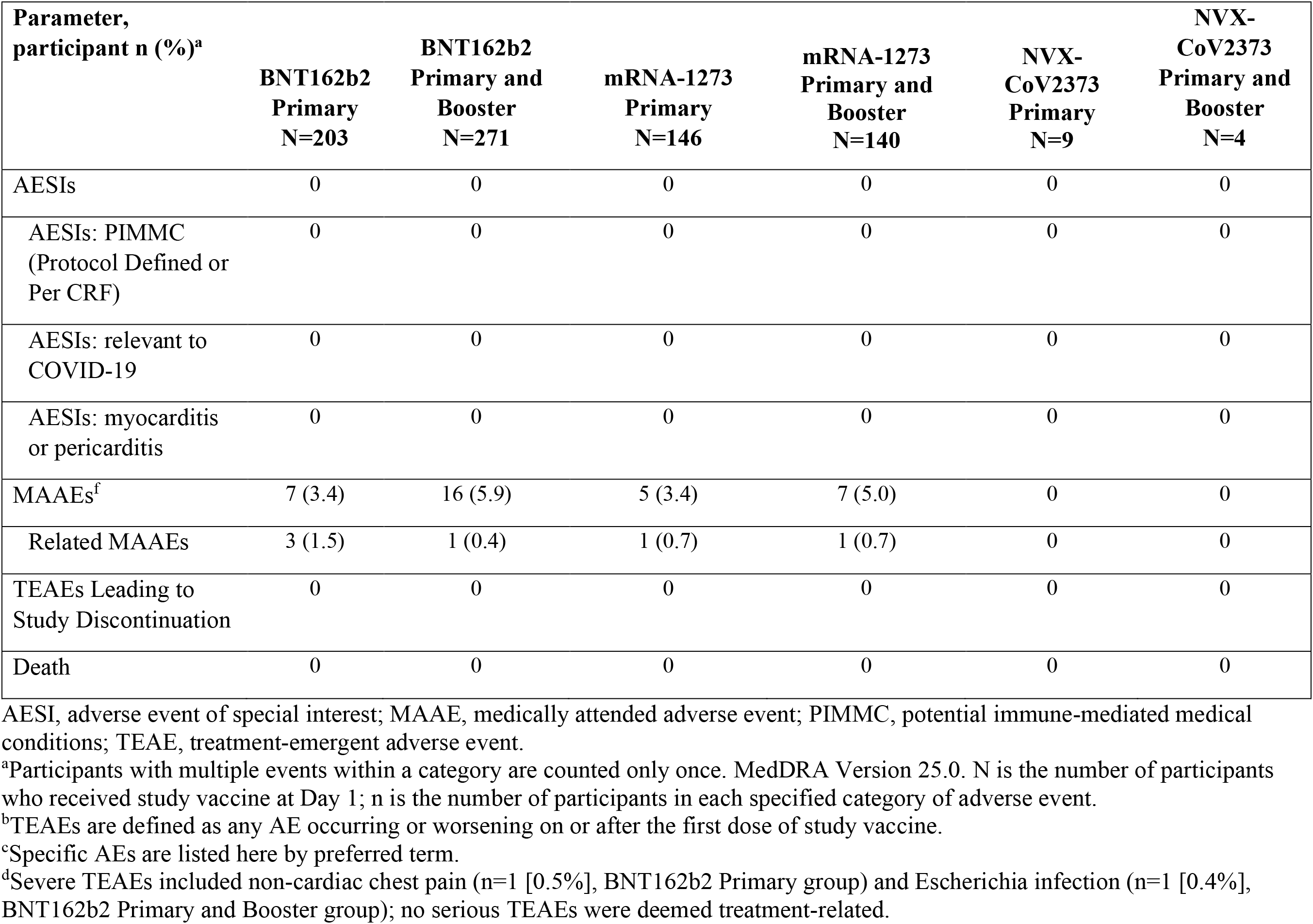
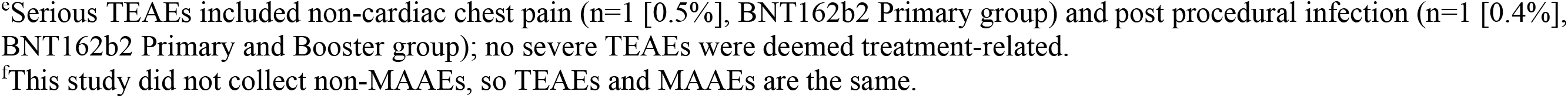
Overall summary of unsolicited TEAEs during the study by prior vaccine type (Safety Analysis Set).

The most common unsolicited AEs and MAAEs were infections and infestations (COVID-19), but these AEs were mild or moderate in severity and not related to treatment. Two participants experienced SAEs (n=1 non-cardiac chest pain, n=1 *Escherichia* infection), but these were unrelated to treatment. No cases of myocarditis or pericarditis were reported (**Table 2, Table S13**).

## 4. Discussion

NVX-CoV2373 was immunogenic and well tolerated when administered as a first or later booster dose after the primary series with or without a first booster of any of the FDA-authorized or approved SARS-CoV-2 vaccines in the US. Immunogenicity was equivalent across three different manufacturing lots, and no new safety signals were identified. NVX-CoV2373 induced robust anti-rS IgG and NAb responses when used either as an initial booster (in participants with prior primary series only) or as a second booster after primary and booster doses. In terms of GMFRs, the improvements in IgG and NAb levels were similar regardless of previous vaccine type, with the greatest fold increase reported in those who had previously received the NVX- CoV2373 vaccine.

Similar improvements in IgG and Nab levels were seen among participants following a homologous prior booster and primary series, with similar post-vaccination GMEUs in all groups, but a trend toward higher responses was observed in previous recipients of the NVX- CoV2373 vaccine. These results are unexpected given recent evidence that, with other vaccine types, heterologous boosting provides better responses than homologous boosting [13–17].

However, the previously published studies focused predominantly on mRNA or other non-protein-based vaccines, with potentially different adaptive immune responses, as compared with the NVX-CoV2373 adjuvanted recombinant protein vaccine evaluated in this study.

SCRs were also similar across the vaccine type groups for IgG and NAbs. Although those with a prior history of receiving NVX-CoV2373 (primary series with or without booster) showed the highest SCRs, the number of participants in this group was small, with wide 95% CIs. For IgG levels in each vaccine type group, SCRs tended to be lower in participants who had a prior heterologous booster when compared with those who had a prior homologous booster or no prior boost, with the exception of recipients of the mRNA-1273 vaccine. These data suggest that NVX-CoV2373-induced IgG seroconversion may be higher when given after a homologous primary and booster regimen. For NAbs, the SCRs were similar between groups, regardless of vaccine type or booster history. Therefore, robust seroconversion responses were also induced in those previously primed with other authorized vaccines when NVX-CoV2373 was administered as a heterologous booster.

Fong et al. recently reported that anti-S IgG levels correlate with vaccine efficacy (VE) estimates for NVX-CoV2373 [18]. In the current study, when IgG levels are converted to BAU/mL (by dividing EU/mL values by 22), IgG levels were above the levels found by Fong et al. in healthy participants who did not subsequently experience a break-through case of COVID-19 (1552 BAU/mL) in all groups in the current study, regardless of prior vaccination history (range: 1669.2 to 14034.0 BAU/mL). Additionally, while all groups showed IgG levels higher than that associated with 87.70% estimated VE (1000 BAU/mL), only those who previously received NVX-CoV2373 had IgG levels above those associated with 94.80% estimated VE (6934 BAU/mL). Therefore, IgG levels seen after boosting with NVX-CoV2373 in the current study are likely to indicate subsequent protection against COVID-19.

Given the recent emergence of immune-evasive SARS-CoV-2 variants such as several of the Omicron lineage [19, 20], IgG cross-reactivity with variants is of critical importance. NVX- CoV2373 induced lower but still robust IgG levels when tested against the Omicron BA.1 and BA.5 subvariants compared with the ancestral strain SARS-CoV-2. This robust broad cross-reactivity was seen regardless of vaccine type received prior to this study, or number of previous doses (primary series with or without homologous booster) prior to the single dose of NVX- CoV2373 in this study. Among those with a primary series only, GMFRs were similar regardless of prior vaccine type. Therefore, NVX-CoV2373 induces robust, broadly cross-reactive humoral responses even against immune-evasive Omicron variants, whether administered as a fully homologous series of NVX-CoV2373 (primary series and first booster) or as a second heterologous booster.

One limitation of the study is that the number of participants who had received prior non-mRNA vaccines (Ad26.COV2.S or NVX-CoV2373) was small. The limited number of participants may have reduced the ability of the study to assess differences in immunogenicity for those groups.

The duration of the study was also relatively short. In addition, the analyses based on prior vaccine type described here were not prespecified analyses, and the study was not necessarily powered to detect differences in these analyses.

## 5. Conclusions

NVX-CoV2373 showed equivalent immunogenicity across different manufacturing lots, as measured by IgG and NAb responses to the vaccine-based ancestral strain. No new safety signals were identified. NVX-CoV2373 was immunogenic regardless of whether it was used as a first booster or later booster dose, and whether it followed earlier doses of NVX-CoV2373 or other authorized vaccines. The broad cross-reactivity of immune response observed with Omicron subvariants suggests that protection afforded by boosting with the ancestral strain-based NVX- CoV2373 vaccine may extend to more recently evolved variants.

## Data Availability

Study information is available at https://clinicaltrials.gov/ct2/show/NCT05463068 and requests will be considered.

## Acknowledgements

Medical writing and editing support were provided by Rebecca Harris, PhD, and Kelly Cameron, PhD of Ashfield MedComms (New York, USA), an Inizio company. The Sponsor acknowledges the contributions and participation of all study volunteers, principal investigators, and investigative site personnel who contributed to the success of the study. The authors would like to thank Kathryn Davis, Kesava Vajjala, and Pratyusha Kajipet for assistance in study conduct, oversight and management of immunogenicity testing.

## Data Sharing Statement

Study information is available at https://clinicaltrials.gov/ct2/show/NCT05463068, and requests will be considered.

## References

[1] Druedahl LC, Minssen T, Price WN. Collaboration in times of crisis: a study on COVID-19 vaccine R&D partnerships. Vaccine 2021;39:6291–5.

[2] Le TT, Andreadakis Z, Kumar A, Román RG, Tollefsen S, Saville M, et al. The COVID-19 vaccine development landscape. Nat Rev Drug Discov 2020;19:305–6.

3. VIPER Group COVID19 Vaccine Tracker Team. COVID19 Vaccine Tracker. 2022. https://covid19.trackvaccines.org/ [accessed 09 February, 2023].

[4] Harvey WT, Carabelli AM, Jackson B, Gupta RK, Thomson EC, Harrison EM, et al. SARS-CoV-2 variants, spike mutations and immune escape. Nat Rev Microbiol 2021;19:409–24.

[5] Jary A, Marot S, Faycal A, Leon S, Sayon S, Zafilaza K, et al. Spike gene evolution and immune escape mutations in patients with mild or moderate forms of COVID-19 and treated with monoclonal antibodies therapies. Viruses 2022;14:226.

[6] Scherer EM, Babiker A, Adelman MW, Allman B, Key A, Kleinhenz JM, et al. SARS-CoV- 2 evolution and immune escape in immunocompromised patients. N Engl J Med 2022;386:2436– 8.

[7] Starr TN, Greaney AJ, Addetia A, Hannon WW, Choudhary MC, Dingens AS, et al. Prospective mapping of viral mutations that escape antibodies used to treat COVID-19. Science 2021;371:850–4.

[8] Parker EPK, Desai S, Marti M, O’Brien KL, Kaslow DC, Kochhar S, et al. Emerging evidence on heterologous COVID-19 vaccine schedules-To mix or not to mix? Lancet Infect Dis 2022;22:438–40.

[9] Siddiqui A, Adnan A, Abbas M, Taseen S, Ochani S, Essar MY. Revival of the heterologous prime-boost technique in COVID-19: an outlook from the history of outbreaks. Health Sci Rep 2022;5:e531.

[10] Dunkle LM, Kotloff KL, Gay CL, Áñez G, Adelglass JM, Barrat Hernández AQ, et al. Efficacy and safety of NVX-CoV2373 in adults in the United States and Mexico. N Engl J Med 2021;386:531–43.

[11] Keech C, Albert G, Cho I, Robertson A, Reed P, Neal S, et al. Phase 1–2 trial of a SARS-CoV-2 recombinant spike protein nanoparticle vaccine. N Engl J Med 2020;383:2320–32.

[12] Formica N, Mallory R, Albert G, Robinson M, Plested JS, Cho I, et al. Different dose regimens of a SARS-CoV-2 recombinant spike protein vaccine (NVX-CoV2373) in younger and older adults: a phase 2 randomized placebo-controlled trial. PLOS Med 2021;18:e1003769.

[13] Atmar RL, Lyke KE, Deming ME, Jackson LA, Branche AR, El Sahly HM, et al. Homologous and heterologous Covid-19 booster vaccinations. N Engl J Med 2022;386:1046–57.

[14] Costa Clemens SA, Weckx L, Clemens R, Almeida Mendes AV, Ramos Souza A, Silveira MBV, et al. Heterologous versus homologous COVID-19 booster vaccination in previous recipients of two doses of CoronaVac COVID-19 vaccine in Brazil (RHH-001): a phase 4, non-inferiority, single blind, randomised study. Lancet (London, England) 2022;399:521–9.

[15] Jara A, Undurraga EA, Zubizarreta JR, González C, Pizarro A, Acevedo J, et al. Effectiveness of homologous and heterologous booster doses for an inactivated SARS-CoV-2 vaccine: a large-scale prospective cohort study. Lancet Glob Health 2022;10:e798–e806.

[16] Mayr FB, Talisa VB, Shaikh O, Yende S, Butt AA. Effectiveness of homologous or heterologous Covid-19 boosters in Veterans. N Engl J Med 2022;386:1375–7.

[17] Tan CS, Collier A-rY, Yu J, Liu J, Chandrashekar A, McMahan K, et al. Durability of heterologous and homologous COVID-19 vaccine boosts. JAMA Netw Open 2022;5:e2226335-e.

[18] Fong Y, Huang Y, Benkeser D, Carpp LN, Áñez G, Woo W, et al. Immune correlates analysis of the PREVENT-19 COVID-19 vaccine efficacy clinical trial. Nature Communications. 2023;14:331.

[19] Liu L, Iketani S, Guo Y, Chan JFW, Wang M, Liu L, et al. Striking antibody evasion manifested by the Omicron variant of SARS-CoV-2. Nature 2022;602:676–81.

[20] Willett BJ, Grove J, MacLean OA, Wilkie C, De Lorenzo G, Furnon W, et al. SARS-CoV-2 Omicron is an immune escape variant with an altered cell entry pathway. Nat Microbiol 2022;7:1161–79.

