## Supplement for "Immunogenicity and safety of NVX-CoV2373 as a homologous or heterologous booster: A phase 3 randomized clinical trial in adults"

### Randomization Methods.

Participants were randomized in a 1:1:1 ratio to receive one intramuscular injection of NVX-CoV-2373 from one of three different manufacturing lots. Randomization was done using an Interactive Web Response System to allocate randomization numbers to individual participants. Both clinical study personnel and participants were blinded to the lot assignment.

**Table S1.** 2019nCoV-307 Study group members (Pubmed Listed, in alphabetical order of institution affiliation).

| Affiliation | Individuals | Location |
|---|---|---|
| Accel Clinical Research | Bruce Rankin | Deland, FL |
| Benchmark Research | Laurence Chu; William Seger; David Ferrera | Austin, TX; Fort Worth, TX; Sacramento, CA |
| Cen/Excel ACMR | Robert Riesenberg | Atlanta, GA |
| CRA Headlands | Jon Finley | Stockbridge, GA |
| CTI | Antoinette Pragalos | Cincinnati, OH |
| Jacksonville Center for Clinical Research | Jeffrey Jacqmein | Jacksonville, FL |
| Long Beach Clinical Trial Services Inc. | Suzanne Fussell, Apinya Vutikullird | Long Beach, CA |
| Lynn Health Science Institute | Carl Griffin | Oklahoma City, OK |
| MedPharmics | Paul Matherne | Gulfport, MS |
| Meridian Clinical Research | Charles Harper; Paul Bradley; David Ensz; Katherine Pearce; Fritz Raiser; Suchet Patel | Norfolk, NE; Savannah, GA; Sioux City, IA; Baton Rouge, LA; Omaha, NE; Endwell, NY |
| Multi-Specialty Research Associates, Inc. | Guy Strauss | Lake City, FL |
| OnSite Clinical Solutions, LLC | Arin Piramzadian | Charlotte, NC |
| Research Centers of America | Howard Schwartz | Hollywood, FL |
| Research Your Health | Jeffrey Adelglass | Plano, TX |
| Rochester Clinical Research | Matthew Davis | Rochester, NY |
| Sundance Clinical Research | Larkin Wadsworth | St. Louis, MO |

| | | |
|---|---|---|
| Tekton Research | Paul Pickrell; Jara McDonald; Robert Lockwood | Austin, TX; San Antonio, TX; Yukon, OK |
| Velocity Clinical Research | David Fried; John Delgado; Margaret Rhee; Marian Shaw | East Greenwich, RI;<br>Grants Pass, OR;<br>Cleveland, OH;<br>Boise, ID |

**Table S2.** Baseline demographics and characteristics (Safety Analysis Set[a]).

| n (%) | Group 1 N=298 | Group 2 N=303 | Group 3 N=304 | Total N=905 |
|---|---|---|---|---|
| **Age (years)[b]** | | | | |
| N | 298 | 303 | 304 | 905 |
| Mean (SD) | 36.2 (8.31) | 36.5 (8.80) | 36.6 (8.06) | 36.4 (8.39) |
| Median | 37.0 | 38.0 | 38.0 | 38.0 |
| Min, Max | (19, 49) | (18, 49) | (18, 49) | (18, 49) |
| **Sex, n (%)** | | | | |
| Male | 123 (41.3) | 138 (45.5) | 118 (38.8) | 379 (41.9) |
| Female | 175 (58.7) | 165 (54.5) | 186 (61.2) | 526 (58.1) |
| **Ethnicity, n (%)** | | | | |
| Hispanic or Latino | 50 (16.8) | 41 (13.5) | 52 (17.1) | 143 (15.8) |
| Not Hispanic or Latino | 245 (82.2) | 259 (85.5) | 248 (81.6) | 752 (83.1) |
| Not Reported/Specified | 3 (1.0) | 3 (1.0) | 4 (1.3) | 10 (1.1) |
| **Race, n (%)** | | | | |
| American Indian or Alaska Native | 3 (1.0) | 1 (0.3) | 3 (1.0) | 7 (0.8) |
| Asian | 11 (3.7) | 10 (3.3) | 9 (3.0) | 30 (3.3) |
| Black or African American | 54 (18.1) | 61 (20.1) | 51 (16.8) | 166 (18.3) |
| Native Hawaiian or Other Pacific Islander | 0 | 0 | 3 (1.0) | 3 (0.3) |
| White | 219 (73.5) | 223 (73.6) | 228 (75.0) | 670 (74.0) |
| Other | 8 (2.7) | 3 (1.0) | 9 (3.0) | 20 (2.2) |
| Not Reported/Specified | 3 (1.0) | 5 (1.7) | 1 (0.3) | 9 (1.0) |
| **BMI, n (%)** | | | | |
| Underweight (<18.5) | 5 (1.7) | 4 (1.3) | 4 (1.3) | 13 (1.4) |
| Healthy (18.5-24.9) | 61 (20.5) | 82 (27.1) | 72 (23.7) | 215 (23.8) |
| Overweight (25.0-29.9) | 82 (27.5) | 86 (28.4) | 64 (21.1) | 232 (25.6) |
| Obese (≥30.0) | 150 (50.3) | 131 (43.2) | 164 (53.9) | 445 (49.2) |
| **Prior or concomitant medications, n (%)** | 204 (68.5) | 196 (64.7) | 220 (72.4) | 620 (68.5) |
| **Time between first dose of previous COVID vaccine and Day 1 dose (weeks)** | | | | |

| | | | | |
|---|---|---|---|---|
| n | 298 | 303 | 304 | 905 |
| Mean (SD) | 66.7 (13.11) | 68.0 (14.12) | 67.7 (12.39) | 67.5 (13.22) |
| Median | 67.7 | 67.6 | 68.3 | 68.0 |
| Min - Max | (26, 102) | (29, 101) | (9, 102) | (9, 102) |
| **Previous SARS-CoV-2 infection, n (%)** | | | | |
| Yes | 17 (5.7) | 13 (4.3) | 14 (4.6) | 44 (4.9) |
| **PCR, n (%)** | | | | |
| Positive | 7 (2.3) | 8 (2.6) | 6 (2.0) | 21 (2.3) |
| Negative | 282 (94.6) | 288 (95.0) | 289 (95.1) | 859 (94.9) |
| Missing | 9 (3.0) | 7 (2.3) | 9 (3.0) | 25 (2.8) |
| **Regimen of previous COVID vaccines received before the study, n (%)** | | | | |
| **NVX-CoV2373** | | | | |
| Primary vaccination series (no booster) | 3 (1.0) | 4 (1.3) | 2 (0.7) | 9 (1.0) |
| Primary series + NVX-CoV2373 booster (homologous boost) | 1 (0.3) | 2 (0.7) | 1 (0.3) | 4 (0.4) |
| Primary series + other booster (heterologous boost) | 4 (1.3) | 4 (1.3) | 4 (1.3) | 12 (1.3) |
| **mRNA-1273** | | | | |
| Primary vaccination series (no booster) | 49 (16.4) | 45 (14.9) | 52 (17.1) | 146 (16.1) |
| Primary series + mRNA-1273 booster (homologous boost) | 51 (17.1) | 46 (15.2) | 43 (14.1) | 140 (15.5) |
| Primary series + other booster (heterologous boost) | 6 (2.0) | 7 (2.3) | 11 (3.6) | 24 (2.7) |
| **BNT162b2** | | | | |
| Primary vaccination series (no booster) | 70 (23.5) | 73 (24.1) | 60 (19.7) | 203 (22.4) |
| Primary series + BNT162b2 booster (homologous boost) | 79 (26.5) | 89 (29.4) | 103 (33.9) | 271 (29.9) |
| Primary series + other booster (heterologous boost) | 12 (4.0) | 13 (4.3) | 15 (4.9) | 40 (4.4) |

| | | | | |
|---|---|---|---|---|
| **Ad26.COV2.S** | | | | |
| Primary vaccination series (no booster) | 7 (2.3) | 7 (2.3) | 8 (2.6) | 22 (2.4) |
| Primary series + Ad26.COV2.S booster (homologous boost) | 4 (1.3) | 1 (0.3) | 1 (0.3) | 6 (0.7) |
| Primary series + other booster (heterologous boost) | 7 (2.3) | 9 (3.0) | 3 (1.0) | 19 (2.1) |
| **Other** | 5 (1.7) | 3 (1.0) | 1 (0.3) | 9 (1.0) |

BMI, body mass index; PCR, polymerase chain reaction; SD, standard deviation.
[a]Safety Analysis Set includes all participants who receive the study vaccine. Treatment group is based on the actual treatment received. N is the number of participants in Safety Analysis Set; n is the number of participants in each specified category.
[b]Age is calculated as the lowest integer result of (Date of Study Day 1 – Date of Birth + 1)/365.25.

**Fig. S1.** IgG ELISA responses and neutralizing antibodies in Per-Protocol Analysis Set before and after boosting with NVX-CoV2373, shown by manufacturing lot.

**(A)**

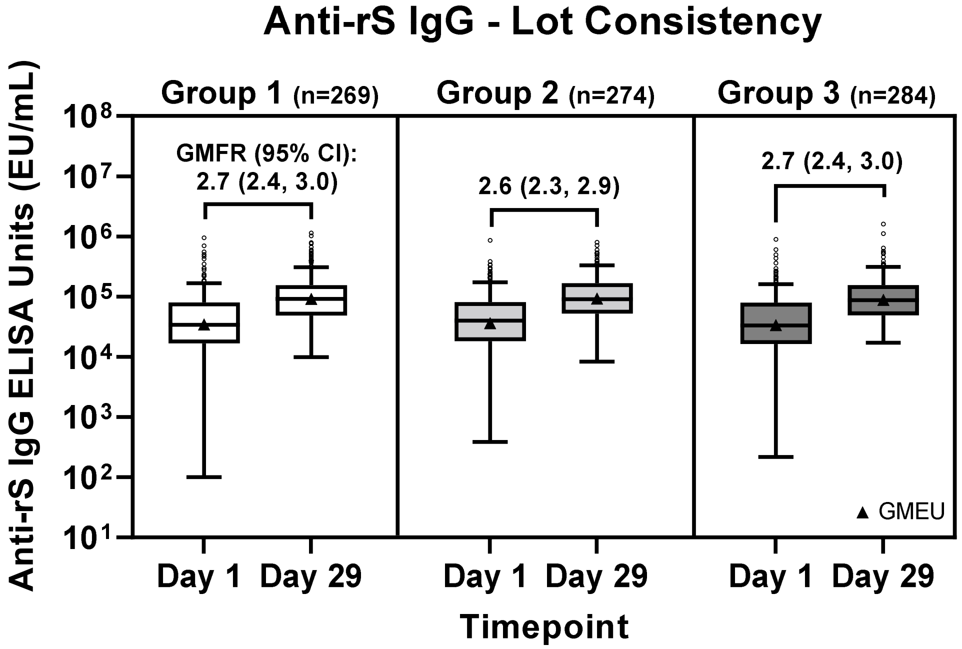


**(B)**

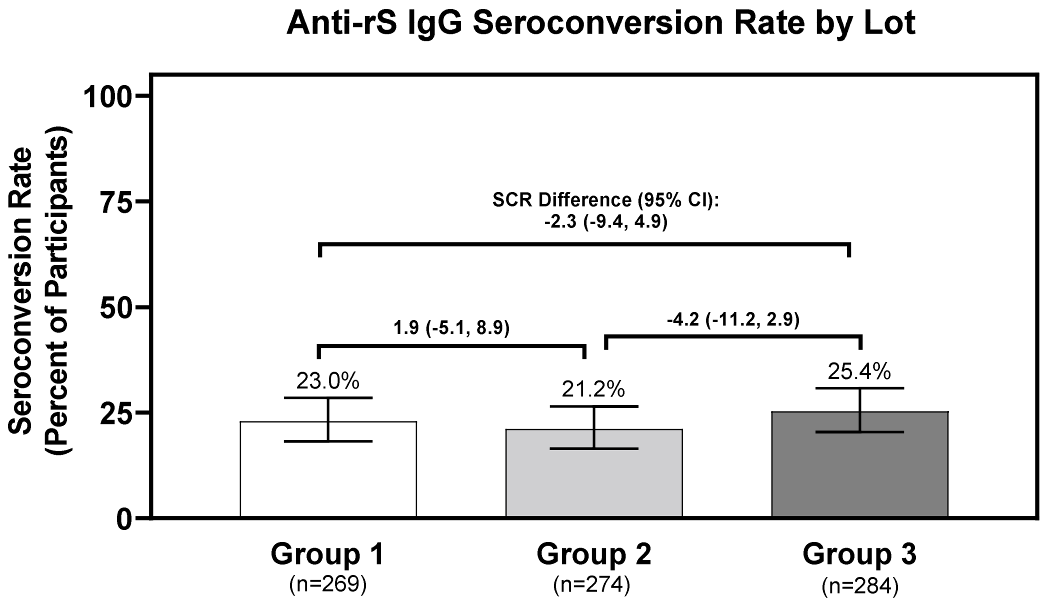

(C)

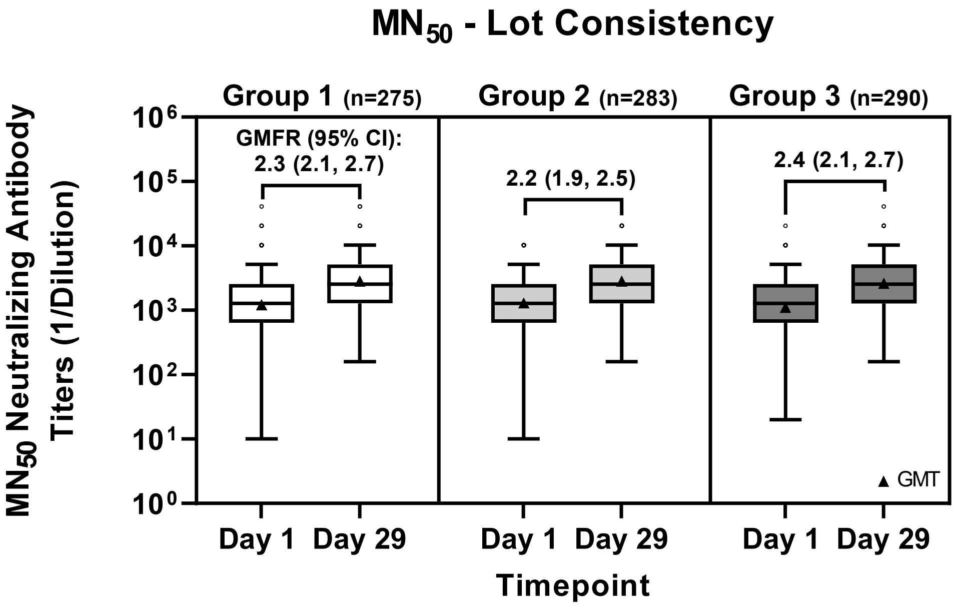


(D)

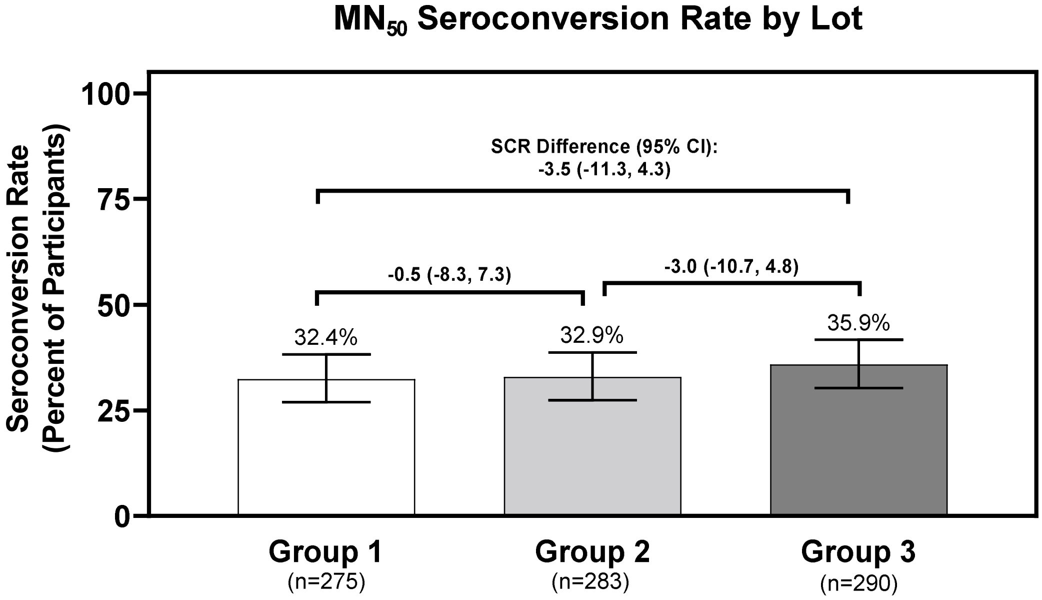


**Fig. S1.** Immunogenicity of NVX-CoV2373 based on anti-rS IgG responses and neutralizing antibodies ($MN_{50}$) was assessed for each of three different manufacturing lots. (A) IgG responses measured by ELISA and (B) seroconversion rates of IgG responses for the three lots are shown. (C) Neutralizing antibody responses measured by microneutralization assay and (D) seroconversion rates of neutralizing antibody responses for the three lots are shown. For IgG and antibody responses, ELISA units or titers are graphed as boxplots with GMEU/GMT as triangles (whiskers drawn using the Tukey method, values outside the whiskers are shown as open circles), and GMFRs with 95% CIs are shown above the boxes. For seroconversion, percentages are graphed with 95% CI, and rate differences with 95% CIs are shown for each lot comparison.

Abbreviations: CI, confidence interval; ELISA, enzyme-linked immunosorbent assay; IgG, immunoglobulin G; MN, microneutralization.

**Table S3.** Geometric mean, geometric mean fold rise, baseline adjusted geometric mean ratio and seroconversion rate of IgG ELISA Units Specific for SARS-CoV-2 rS, by manufacturing lot (Per-Protocol Analysis Set).

| **Parameter**[a] | **Group 1 N=275** | **Group 2 N=283** | **Group 3 N=290** | **Total N=848** |
|---|---|---|---|---|
| Baseline (Day 1) | | | | |
| N | 269 | 274 | 284 | 827 |
| GMEU | 34294.3 | 36142.6 | 33233.4 | 34521.4 |
| 95% CI | (29549.6, 39800.9) | (31430.8, 41560.8) | (28863.0, 38265.4) | (31792.6, 37484.5) |
| Day 29 | | | | |
| N | 269 | 274 | 284 | 827 |
| GMEU | 91945.8 | 93084.2 | 88811.3 | 91228.2 |
| 95% CI | (82821.7, 102075.0) | (84250.8, 102843.7) | (80851.9, 97554.2) | (86164.5, 96589.6) |
| Day 29 vs Baseline | | | | |
| N | 269 | 274 | 284 | 827 |
| GMFR | 2.7 | 2.6 | 2.7 | 2.6 |
| 95% CI | (2.4, 3.0) | (2.3, 2.9) | (2.4, 3.0) | (2.5, 2.8) |
| GMEUR[b] | | Group 1 vs. Group 2 | Group 1 vs. Group 3 | |
| Estimate | | 1.01 | 1.02 | |
| 95% CI | | (0.90, 1.13) | (0.91, 1.15) | |
| | | | Group 2 vs. Group 3 | |
| | | | 1.01 | |
| | | | (0.91, 1.13) | |

| | | | | |
|---|---|---|---|---|
| Day 29 | | | | |
| N | 269 | 274 | 284 | 827 |
| SCR[c] | 62 ( 23.0) | 58 ( 21.2) | 72 ( 25.4) | 192 ( 23.2) |
| 95% CI | (18.2, 28.5) | (16.5, 26.5) | (20.4, 30.8) | (20.4, 26.2) |
| | | Group 1 vs. Group 2 | Group 1 vs. Group 3 | |
| SCRs Diff | | 1.9 | -2.3 | |
| 95% CI | | (-5.1, 8.9) | (-9.4, 4.9) | |
| | | | Group 2 vs. Group 3 | |
| | | | -4.2 | |
| | | | (-11.2, 2.9) | |

GMEU, geometric mean ELISA unit; GMEUR, geometric mean ELISA unit ratio (between groups); GMFR, geometric mean fold rise (Day 29 / Baseline within groups); SCR, seroconversion rate; CI, confidence interval; N, number of participants in Per Protocol Analysis Set; n, number of participants with non-missing results at each visit.

[a]Individual antibody values recorded as below the lower limit of quantitation (LLoQ) will be set to half LLoQ.

[b]GMEUR is obtained from a mixed effects model with study vaccine group and baseline IgG ELISA unit as covariate. The ratios of geometric least square (LS) means and 95% confidence intervals for the ratios are calculated by back transforming mean differences and 95% confidence limits for the differences of log transformed IgG ELISA units between two vaccine groups.

[c]SCR is defined as the percentage of participants with post vaccination IgG ELISA ≥ 4 times the baseline value; Percentages are based on the number of participants with non-missing IgG ELISA units in Per Protocol Analysis Set. Clopper-Pearson method is applied to calculate CIs of SCR for each group. CIs of the difference in SCRs for paired comparisons is based on the Miettinen and Nurminen method.

**Table S4.** Summary of geometric mean, geometric mean fold rise and baseline adjusted geometric mean ratio of neutralizing antibody titers for SARS-CoV-2 wildtype virus, by manufacturing lot (Per Protocol Analysis Set).

| **Parameter[a]** | **Group 1 N=275** | **Group 2 N=283** | **Group 3 N=290** | **Total N=848** |
|---|---|---|---|---|
| **Baseline (Day 1)** | | | | |
| **N** | 275 | 283 | 290 | 848 |
| **GMT** | 1211.0 | 1289.4 | 1103.7 | 1198.0 |
| **95% CI** | (1028.9, 1425.2) | (1114.2, 1492.3) | (950.1, 1282.1) | (1097.0, 1308.2) |
| **Day 29** | | | | |
| **N** | 275 | 283 | 290 | 848 |
| **GMT** | 2838.7 | 2823.5 | 2603.2 | 2750.9 |
| **95% CI** | (2532.6, 3181.8) | (2529.0, 3152.4) | (2341.2, 2894.6) | (2582.1, 2930.8) |
| **Day 29 vs Baseline** | | | | |
| **n** | 275 | 283 | 290 | 848 |
| **GMFR** | 2.3 | 2.2 | 2.4 | 2.3 |
| **95% CI** | (2.1, 2.7) | (1.9, 2.5) | (2.1, 2.7) | (2.1, 2.5) |
| **GMTR[b]** | | Group 1 vs. Group 2 | Group 1 vs. Group 3 | |
| **Estimate** | | 1.03 | 1.05 | |
| **95% CI** | | (0.91, 1.17) | (0.93, 1.19) | |
| | | | Group 2 vs. Group 3 | |
| | | | 1.02 | |
| | | | (0.90, 1.15) | |
| **Day 29** | | | | |
| **n** | 275 | 283 | 290 | 848 |

| Parameter[a] | Group 1 N=275 | Group 2 N=283 | Group 3 N=290 | Total N=848 |
|---|---|---|---|---|
| **SCR[c]** | 89 ( 32.4) | 93 ( 32.9) | 104 ( 35.9) | 286 ( 33.7) |
| **95% CI** | (26.9, 38.2) | (27.4, 38.7) | (30.3, 41.7) | (30.5, 37.0) |
| | | Group 1 vs. Group 2 | Group 1 vs. Group 3 | |
| **SCRs Diff** | | -0.5 | -3.5 | |
| **95% CI** | | (-8.3, 7.3) | (-11.3, 4.3) | |
| | | | Group 2 vs. Group 3 | |
| | | | -3.0 | |
| | | | (-10.7, 4.8) | |

GMFR, geometric mean fold rise (Day 29 / Baseline within groups); GMT, geometric mean titer; GMTR, geometric mean titer ratio (between groups); SCR, seroconversion rate; CI, confidence interval; N, number of participants in Per Protocol Analysis Set; n, number of participants with non-missing results at each visit.

[a]Individual antibody values recorded as below the lower limit of quantitation (LLoQ) will be set to half LLoQ.

[b]GMTR is obtained from a mixed effects model with study vaccine group and baseline antibody titer as covariate. The ratios of geometric least square (LS) means and 95% confidence intervals for the ratios are calculated by back transforming mean differences and 95% confidence limits for the differences of log transformed antibody titers between two vaccine groups.

[c]SCR is defined as the percentage of participants with post vaccination MN50 titers ≥ 4 times the baseline value; Percentages are based on the number of participants with non-missing IgG ELISA units in Per Protocol Analysis Set. Clopper-Pearson method is applied to calculate CIs of SCR for each group. CIs of the difference in SCRs for paired comparisons is based on the Miettinen and Nurminen method.

**Table S5.** Summary of geometric mean, geometric mean fold rise, baseline adjusted geometric mean ratio, and seroconversion rate of IgG ELISA units specific for SARS-CoV-2 rS (no prior booster).

| Parameter[a] | BNT162b Primary Series N=190 | mRNA-1273 Primary Series N=131 | Ad26.COV2.S Primary Series N=19 | NVX-CoV2373 Primary Series N=7 |
|---|---|---|---|---|
| Baseline (Day 1) | | | | |
| n | 187 | 125 | 19 | 7 |
| GMEU | 24487.8 | 32717.2 | 12632.2 | 33086.9 |
| 95% CI | (20577.6, 29141.1) | (25419.5, 42110.1) | (7995.0, 19959.0) | (7807.0, 140225.4) |
| Day 29 | | | | |
| n | 187 | 125 | 19 | 7 |
| GMEU | 75135.7 | 88669.9 | 47455.9 | 263387.4 |
| 95% CI | (66853.4, 84444.1) | (75395.8, 104281.1) | (35971.4, 62607.1) | (187909.7, 369182.3) |
| Day 29 vs Baseline | | | | |
| n | 187 | 125 | 19 | **7** |
| GMFR | 3.1 | 2.7 | 3.8 | 8.0 |
| 95% CI | (2.6, 3.6) | (2.2, 3.3) | (2.5, 5.7) | (1.7, 37.1) |
| Day 29 | | | | |
| n | 187 | 125 | 19 | 7 |
| SCR[b] | 49 ( 26.2) | 30 ( 24.0) | 8 ( 42.1) | 4 ( 57.1) |
| 95% CI | (20.1, 33.1) | (16.8, 32.5) | (20.3, 66.5) | (18.4, 90.1) |

GMEU, geometric mean ELISA unit; GMFR, geometric mean fold rise (Day 29 / Baseline within groups); SCR, seroconversion rate; CI, confidence interval; N, number of participants in Per Protocol Analysis Set; n, number of participants with non-missing results at each visit.
[a]Individual antibody values recorded as below the lower limit of quantitation (LLoQ) will be set to half LLoQ.
[b]SCR is defined as the percentage of participants with post vaccination IgG ELISA ≥ 4 times the baseline value; Percentages are based on the number of participants with non-missing IgG ELISA units in Per Protocol Analysis Set. Clopper-Pearson method is applied to calculate CIs of

SCR for each group. CIs of the difference in SCRs for paired comparisons is based on the Miettinen and Nurminen method.

**Table S6.** Summary of geometric mean, geometric mean fold rise, baseline adjusted geometric mean ratio, and seroconversion rate of neutralizing antibody titers for SARS-CoV-2 wildtype virus (no prior booster).

| Parameter[a] | BNT162b Primary Series N=190 | mRNA-1273 Primary Series N=131 | Ad26.COV2.S Primary Series N=19 | NVX-CoV2373 Primary Series N=7 |
|---|---|---|---|---|
| Baseline (Day 1) | | | | |
| n | 190 | 131 | 19 | 7 |
| GMT | 918.4 | 1226.9 | 740.6 | 1050.0 |
| 95% CI | (753.0, 1120.1) | (956.5, 1573.9) | (458.2, 1196.8) | (253.9, 4342.9) |
| Day 29 | | | | |
| n | 190 | 131 | 19 | 7 |
| GMT | 2486.4 | 2906.6 | 1983.1 | 4200.1 |
| 95% CI | (2190.8, 2821.8) | (2430.7, 3475.8) | (1365.8, 2879.2) | (2282.7, 7728.2) |
| Day 29 vs Baseline | | | | |
| n | 190 | 131 | 19 | 7 |
| GMFR | 2.7 | 2.4 | 2.7 | 4.0 |
| 95% CI | (2.3, 3.2) | (1.9, 2.9) | (1.6, 4.4) | (1.0, 16.0) |
| Day 29 | | | | |
| n | 190 | 131 | 19 | 7 |
| SCR[b] | 70 ( 36.8) | 42 ( 32.1) | 8 ( 42.1) | 3 ( 42.9) |
| 95% CI | (30.0, 44.1) | (24.2, 40.8) | (20.3, 66.5) | (9.9, 81.6) |

GMFR, geometric mean fold rise (Day 29 / Baseline within groups); GMT, geometric mean titer; SCR, seroconversion rate; CI, confidence interval; N, number of participants in Per Protocol Analysis Set; n, number of participants with non-missing results at each visit.

[a]Individual antibody values recorded as below the lower limit of quantitation (LLoQ) will be set to half LLoQ.

[b]SCR is defined as the percentage of participants with post vaccination MN50 titers ≥ 4 times the baseline value; Percentages are based on the number of participants with non-missing IgG ELISA units in Per Protocol Analysis Set. Clopper-Pearson method is applied to calculate CIs of SCR for each group. CIs of the difference in SCRs for paired comparisons is based on the Miettinen and Nurminen method.

**Table S7.** Summary of geometric mean, geometric mean fold rise, baseline adjusted geometric mean ratio, and seroconversion rate of IgG ELISA units specific for SARS-CoV-2 rS (homologous primary series and prior booster).

| Parameter[a] | BNT162b Primary Series + Booster N=260 | mRNA-1273 Primary Series + Booster N=134 | Ad26.COV2.S Primary Series + Booster N=6 | NVX-CoV2373 Primary Series + Booster N=4 |
|---|---|---|---|---|
| Baseline (Day 1) | | | | |
| n | 254 | 130 | 6 | 4 |
| GMEU | 35622.4 | 57835.8 | 17724.5 | 45521.4 |
| 95% CI | (31105.3, 40795.4) | (48687.0, 68703.8) | (1994.9, 157480.7) | (1784.0, 1161528.4) |
| Day 29 | | | | |
| n | 254 | 130 | 6 | 4 |
| GMEU | 91465.3 | 116472.6 | 93596.5 | 294763.2 |
| 95% CI | (83279.9, 100455.3) | (100957.3, 134372.2) | (38821.0, 225659.0) | (66014.8, 1316148.7) |
| Day 29 vs Baseline | | | | |
| n | 254 | 130 | 6 | 4 |
| GMFR | 2.6 | 2.0 | 5.3 | 6.5 |
| 95% CI | (2.3, 2.8) | (1.8, 2.3) | (1.2, 24.1) | (0.1, 661.3) |
| Day 29 | | | | |
| n | 254 | 130 | 6 | 4 |
| SCR[b] | 68 ( 26.8) | 16 ( 12.3) | 2 ( 33.3) | 2 ( 50.0) |
| 95% CI | (21.4, 32.7) | (7.2, 19.2) | (4.3, 77.7) | (6.8, 93.2) |

GMEU, geometric mean ELISA unit; GMFR, geometric mean fold rise (Day 29 / Baseline within groups); SCR, seroconversion rate; CI, confidence interval; N, number of participants in Per Protocol Analysis Set; n, number of participants with non-missing results at each visit.
[a]Individual antibody values recorded as below the lower limit of quantitation (LLoQ) will be set to half LLoQ.
[b]SCR is defined as the percentage of participants with post vaccination IgG ELISA ≥ 4 times the baseline value; Percentages are based on the number of participants with non-missing IgG ELISA units in Per Protocol Analysis Set. Clopper-Pearson method is applied to calculate CIs of

SCR for each group. CIs of the difference in SCRs for paired comparisons is based on the Miettinen and Nurminen method.

**Table S8.** Summary of geometric mean, geometric mean fold rise, baseline adjusted geometric mean ratio, and seroconversion rate of IgG ELISA units specific for SARS-CoV-2 rS (heterologous primary series and prior booster).

| Parameter[a] | BNT162b Primary Series + Other Booster N=37 | mRNA-1273 Primary Series + Other Booster N=24 | Ad26.COV2.S Primary Series + Other Booster N=18 | NVX-CoV2373 Primary Series + Other Booster N=12 |
|---|---|---|---|---|
| Baseline (Day 1) | | | | |
| n | 35 | 24 | 18 | 12 |
| GMEU | 40782.5 | 40649.9 | 21727.4 | 141217.8 |
| 95% CI | (29491.1, 56397.0) | (25046.5, 65974.0) | (15555.9, 30347.2) | (100615.4, 198204.8) |
| Day 29 | | | | |
| n | 35 | 24 | 18 | 12 |
| GMEU | 80799.0 | 105222.4 | 70966.2 | 308747.2 |
| 95% CI | (61426.2, 106281.5) | (74241.1, 149132.3) | (50212.3, 100298.2) | (225871.1, 422031.9) |
| Day 29 vs Baseline | | | | |
| n | 35 | 24 | 18 | 12 |
| GMFR | 2.0 | 2.6 | 3.3 | 2.2 |
| 95% CI | (1.6, 2.4) | (1.9, 3.5) | (2.1, 5.1) | (1.5, 3.1) |
| Day 29 | | | | |
| n | 35 | 24 | 18 | 12 |
| SCR[b] | 4 ( 11.4) | 4 ( 16.7) | 4 ( 22.2) | 1 ( 8.3) |
| 95% CI | (3.2, 26.7) | (4.7, 37.4) | (6.4, 47.6) | (0.2, 38.5) |

GMEU, geometric mean ELISA unit; GMFR, geometric mean fold rise (Day 29 / Baseline within groups); SCR, seroconversion rate; CI, confidence interval; N, number of participants in Per Protocol Analysis Set; n, number of participants with non-missing results at each visit.
[a]Individual antibody values recorded as below the lower limit of quantitation (LLoQ) will be set to half LLoQ.
[b]SCR is defined as the percentage of participants with post vaccination IgG ELISA ≥ 4 times the baseline value; Percentages are based on the number of participants with non-missing IgG

ELISA units in Per Protocol Analysis Set. Clopper-Pearson method is applied to calculate CIs of SCR for each group. CIs of the difference in SCRs for paired comparisons is based on the Miettinen and Nurminen method.

**Table S9.** Summary of geometric mean, geometric mean fold rise, baseline adjusted geometric mean ratio, and seroconversion rate of neutralizing antibody titers for SARS-CoV-2 wildtype virus (homologous primary series and prior booster).

| Parameter[a] | BNT162b Primary Series + Booster N=260 | mRNA-1273 Primary Series + Booster N=134 | Ad26.COV2.S Primary Series + Booster N=6 | NVX-CoV2373 Primary Series + Booster N=4 |
|---|---|---|---|---|
| Baseline (Day 1) | | | | |
| n | 260 | 134 | 6 | 4 |
| GMT | 1111.3 | 1754.9 | 905.1 | 1810.2 |
| 95% CI | (955.2, 1293.0) | (1442.1, 2135.4) | (117.2, 6992.4) | (58.7, 55848.9) |
| Day 29 | | | | |
| n | 260 | 134 | 6 | 4 |
| GMT | 2499.3 | 3264.6 | 4063.7 | 12177.5 |
| 95% CI | (2236.4, 2793.2) | (2771.2, 3845.7) | (1238.9, 13329.3) | (3039.6, 48785.8) |
| Day 29 vs Baseline | | | | |
| n | 260 | 134 | 6 | 4 |
| GMFR | 2.2 | 1.9 | 4.5 | 6.7 |
| 95% CI | (2.0, 2.5) | (1.6, 2.1) | (0.7, 29.0) | (0.1, 686.2) |
| Day 29 | | | | |
| n | 260 | 134 | 6 | 4 |
| SCR[b] | 96 ( 36.9) | 36 ( 26.9) | 2 ( 33.3) | 1 ( 25.0) |
| 95% CI | (31.0, 43.1) | (19.6, 35.2) | (4.3, 77.7) | (0.6, 80.6) |

GMFR, geometric mean fold rise (Day 29 / Baseline within groups); GMT, geometric mean titer; SCR, seroconversion rate; CI, confidence interval; N, number of participants in Per Protocol Analysis Set; n, number of participants with non-missing results at each visit.

[a]Individual antibody values recorded as below the lower limit of quantitation (LLoQ) will be set to half LLoQ.

[b]SCR is defined as the percentage of participants with post vaccination MN50 titers ≥ 4 times the baseline value; Percentages are based on the number of participants with non-missing IgG ELISA units in Per Protocol Analysis Set. Clopper-Pearson method is applied to calculate CIs of

SCR for each group. CIs of the difference in SCRs for paired comparisons is based on the Miettinen and Nurminen method.

**Table S10.** Summary of geometric mean, geometric mean fold rise, baseline adjusted geometric mean ratio, and seroconversion rate of neutralizing antibody titers for SARS-CoV-2 wildtype virus (heterologous primary series and prior booster).

| Parameter[a] | BNT162b Primary Series + Other Booster N=37 | mRNA-1273 Primary Series + Other Booster N=24 | Ad26.COV2.S Primary Series + Other Booster N=18 | NVX-CoV2373 Primary Series + Other Booster N=12 |
|---|---|---|---|---|
| Baseline (Day 1) | | | | |
| N | 37 | 24 | 18 | 12 |
| GMT | 1487.0 | 1140.4 | 1097.3 | 6088.7 |
| 95% CI | (1019.0, 2169.8) | (691.1, 1881.6) | (721.7, 1668.2) | (4368.7, 8485.9) |
| Day 29 | | | | |
| N | 37 | 24 | 18 | 12 |
| GMT | 2759.2 | 2712.2 | 2463.3 | 9122.8 |
| 95% CI | (2115.5, 3598.8) | (1810.7, 4062.6) | (1622.5, 3739.8) | (6650.4, 12514.3) |
| Day 29 vs Baseline | | | | |
| N | 37 | 24 | 18 | 12 |
| GMFR | 1.9 | 2.4 | 2.2 | 1.5 |
| 95% CI | (1.4, 2.5) | (1.6, 3.5) | (1.4, 3.6) | (1.0, 2.3) |
| Day 29 | | | | |
| N | 37 | 24 | 18 | 12 |
| SCR[b] | 13 ( 35.1) | 8 ( 33.3) | 5 ( 27.8) | 1 ( 8.3) |
| 95% CI | (20.2, 52.5) | (15.6, 55.3) | (9.7, 53.5) | (0.2, 38.5) |

GMFR, geometric mean fold rise (Day 29 / Baseline within groups); GMT, geometric mean titer; SCR, seroconversion rate; CI, confidence interval; N, number of participants in Per Protocol Analysis Set; n, number of participants with non-missing results at each visit.
[a]Individual antibody values recorded as below the lower limit of quantitation (LLoQ) will be set to half LLoQ.
[b]SCR is defined as the percentage of participants with post vaccination MN50 titers ≥ 4 times the baseline value; Percentages are based on the number of participants with non-missing IgG ELISA units in Per Protocol Analysis Set. Clopper-Pearson method is applied to calculate CIs of

SCR for each group. CIs of the difference in SCRs for paired comparisons is based on the Miettinen and Nurminen method.

.

**Table S11.** Summary of geometric mean, geometric mean fold rise, and seroconversion rate of IgG ELISA units specific for variant SARS-CoV-2 rS (no prior booster).

| Parameter[a] | Ancestral | | | Omicron BA.1 | | | Omicron BA.5 | | |
|---|---|---|---|---|---|---|---|---|---|
| | **mRNA-1273** | **BNT162b2** | **NVX-CoV2373** | **mRNA-1273** | **BNT162b2** | **NVX-CoV2373** | **mRNA-1273** | **BNT162b2** | **NVX-CoV2373** |
| Baseline (Day 1) | | | | | | | | | |
| n | 40 | 64 | 7 | 40 | 64 | 7 | 40 | 64 | 7 |
| GMEU | 26307.7 | 26321.8 | 33086.9 | 13163.2 | 11778.9 | 15974.0 | 9909.2 | 9606.6 | 13719.8 |
| 95% CI | (15649.7, 44224.2) | (19153.0, 36173.9) | (7807.0, 140225.4) | (8345.7, 20761.7) | (8526.2, 16272.6) | (3621.7, 70455.3) | (6023.4, 16301.7) | (6993.5, 13196.2) | (3050.0, 61715.5) |
| Day 29 | | | | | | | | | |
| n | 40 | 64 | 7 | 40 | 64 | 7 | 40 | 64 | 7 |
| GMEU | 96562.4 | 82897.0 | 263387.4 | 54965.3 | 41935.9 | 161257.6 | 44533.9 | 36721.1 | 127912.8 |
| 95% CI | (70145.2, 132928.5) | (67445.2, 101888.8) | (187909.7, 369182.3) | (39393.3, 76692.9) | (33480.6, 52526.6) | (101349.9, 256576.6) | (32005.1, 61967.3) | (29716.4, 45376.8) | (83403.7, 196174.4) |
| Day 29 vs Baseline | | | | | | | | | |
| n | 40 | 64 | 7 | 40 | 64 | 7 | 40 | 64 | 7 |
| GMFR | 3.7 | 3.1 | 8.0 | 4.2 | 3.6 | 10.1 | 4.5 | 3.8 | 9.3 |
| 95% CI | (2.3, 5.8) | (2.3, 4.3) | (1.7, 37.1) | (2.6, 6.6) | (2.6, 4.8) | (2.2, 47.4) | (2.9, 7.0) | (2.8, 5.2) | (2.2, 39.3) |
| Day 29 | | | | | | | | | |
| n | 40 | 64 | 7 | 40 | 64 | 7 | 40 | 64 | 7 |
| SCR[b] | 15 ( 37.5) | 18 ( 28.1) | 4 ( 57.1) | 18 ( 45.0) | 24 ( 37.5) | 5 ( 71.4) | 19 ( 47.5) | 24 ( 37.5) | 5 ( 71.4) |
| 95% CI | (22.7, 54.2) | (17.6, 40.8) | (18.4, 90.1) | (29.3, 61.5) | (25.7, 50.5) | (29.0, 96.3) | (31.5, 63.9) | (25.7, 50.5) | (29.0, 96.3) |

GMEU, geometric mean ELISA unit; GMFR, geometric mean fold rise (Day 29 / Baseline within groups); SCR, seroconversion rate; CI, confidence interval; N, number of participants in Per Protocol Analysis Set; n, number of participants with non-missing results at each visit.

[a]Individual antibody values recorded as below the lower limit of quantitation (LLoQ) will be set to half LLoQ.

[b]SCR is defined as the percentage of participants with post vaccination IgG ELISA ≥ 4 times the baseline value; Percentages are based on the number of participants with non-missing IgG ELISA units in Per Protocol Analysis Set. Clopper-Pearson method is applied to calculate CIs of SCR for each group. CIs of the difference in SCRs for paired comparisons is based on the Miettinen and Nurminen method.

**Table S12.** Summary of geometric mean, geometric mean fold rise, and seroconversion rate of IgG ELISA units specific for variant SARS-CoV-2 rS (homologous primary series and prior booster).

| Parameter[a] | Ancestral | | Omicron BA.1 | | Omicron BA.5 | |
|---|---|---|---|---|---|---|
| | **mRNA-1273** | **BNT162b2** | **mRNA-1273** | **BNT162b2** | **mRNA-1273** | **BNT162b2** |
| Baseline (Day 1) | | | | | | |
| n | 60 | 60 | 60 | 59 | 60 | 59 |
| GMEU | 56256.2 | 32892.6 | 24253.9 | 13541.2 | 19862.4 | 10733.0 |
| 95% CI | (43589.9, 72603.0) | (24828.7, 43575.4) | (18720.6, 31422.7) | (10133.8, 18094.3) | (15401.4, 25615.7) | (8119.3, 14187.9) |
| Day 29 | | | | | | |
| n | 60 | 60 | 59 | 60 | 59 | 60 |
| GMEU | 117410.6 | 85188.6 | 52503.7 | 39696.8 | 43471.3 | 33938.2 |
| 95% CI | (94291.6, 146198.1) | (71045.3, 102147.6) | (42403.6, 65009.5) | (32975.2, 47788.5) | (34794.5, 54311.9) | (27704.0, 41575.3) |
| Day 29 vs Baseline | | | | | | |
| n | 60 | 60 | 59 | 59 | 59 | 59 |
| GMFR | 2.1 | 2.6 | 2.2 | 2.9 | 2.2 | 3.2 |
| 95% CI | (1.7, 2.6) | (2.1, 3.2) | (1.8, 2.8) | (2.4, 3.6) | (1.8, 2.8) | (2.6, 3.9) |
| Day 29 | | | | | | |
| n | 60 | 60 | 59 | 59 | 59 | 59 |
| SCR[b] | 10 ( 16.7) | 15 ( 25.0) | 10 ( 16.9) | 18 ( 30.5) | 13 ( 22.0) | 18 ( 30.5) |
| 95% CI | (8.3, 28.5) | (14.7, 37.9) | (8.4, 29.0) | (19.2, 43.9) | (12.3, 34.7) | (19.2, 43.9) |

GMEU, geometric mean ELISA unit; GMFR, geometric mean fold rise (Day 29 / Baseline within groups); SCR, seroconversion rate; CI, confidence interval; N, number of participants in Per Protocol Analysis Set; n, number of participants with non-missing results at each visit.

[a]Individual antibody values recorded as below the lower limit of quantitation (LLoQ) will be set to half LLoQ.

[b]SCR is defined as the percentage of participants with post vaccination IgG ELISA ≥ 4 times the baseline value; Percentages are based on the number of participants with non-missing IgG ELISA units in Per Protocol Analysis Set. Clopper-Pearson method is applied to calculate CIs of SCR for each group. CIs of the difference in SCRs for paired comparisons is based on the Miettinen and Nurminen method.

**Table S13.** Overall summary of unsolicited TEAEs during the study by manufacturing lot (Safety Analysis Set).

| **Parameter, participant n (%)[a]** | **Group 1 N=298** | **Group 2 N=303** | **Group 3 N=304** | **Overall N=905** |
|---|---|---|---|---|
| TEAEs[b] | 15 (5.0) | 12 (4.0) | 12 (3.9) | 39 (4.3) |
| Related TEAEs[c] | 2 (0.7) | 2 (0.7) | 2 (0.7) | 6 (0.7) |
| Headache | 0 | 1 (0.3) | 0 | 1 (0.1) |
| Presyncope | 0 | 0 | 1 (0.3) | 1 (0.1) |
| Nausea | 0 | 1 (0.3) | 1 (0.3) | 2 (0.2) |
| Diarrhea | 0 | 0 | 1 (0.3) | 1 (0.1) |
| Urticaria | 0 | 1 (0.3) | 0 | 1 (0.1) |
| Body temperature fluctuation | 0 | 0 | 1 (0.3) | 1 (0.1) |
| Lymphadenopathy | 1 (0.3) | 0 | 0 | 1 (0.1) |
| Myofascial pain syndrome | 1 (0.3) | 0 | 0 | 1 (0.1) |
| Hot flush | 0 | 1 (0.3) | 0 | 1 (0.1) |
| Severe TEAEs | 0 | 1 (0.3) | 1 (0.3) | 2 (0.2) |
| Post procedural infection | 0 | 0 | 1 (0.3) | 1 (0.1) |
| Non-cardiac chest pain | 0 | 1 (0.3) | 0 | 1 (0.1) |
| Severe Related TEAEs | 0 | 0 | 0 | 0 |
| Serious TEAEs | 0 | 1 (0.3) | 1 (0.3) | 2 (0.2) |
| Escherichia infection | 0 | 0 | 1 (0.3) | 1 (0.1) |
| Non-cardiac chest pain | 0 | 1 (0.3) | 0 | 1 (0.1) |
| Serious Related TEAEs | 0 | 0 | 0 | 0 |
| AESIs | 0 | 0 | 0 | 0 |

| **Parameter, participant n (%)[a]** | **Group 1 N=298** | **Group 2 N=303** | **Group 3 N=304** | **Overall N=905** |
|---|---|---|---|---|
| AESIs: PIMMC (Protocol Defined or Per CRF) | 0 | 0 | 0 | 0 |
| AESIs: relevant to COVID-19 | 0 | 0 | 0 | 0 |
| AESIs: myocarditis or pericarditis | 0 | 0 | 0 | 0 |
| MAAEs[d] | 15 (5.0) | 12 (4.0) | 12 (3.9) | 39 (4.3) |
| Related MAAEs | 2 (0.7) | 2 (0.7) | 2 (0.7) | 6 (0.7) |
| TEAEs Leading to Study Discontinuation | 0 | 0 | 0 | 0 |
| Death | 0 | 0 | 0 | 0 |

AESI, adverse event of special interest; MAAE, medically attended adverse event; PIMMC, potential immune-mediated medical conditions; TEAE, treatment-emergent adverse event.

[a]Participants with multiple events within a category are counted only once. MedDRA Version 25.0. N is the number of participants who received study vaccine at Day 1; n is the number of participants in each specified category of adverse event.

[b]TEAEs are defined as any AE occurring or worsening on or after the first dose of study vaccine.

[c]Specific AEs are listed here by preferred term.

[d]This study did not collect non-MAAEs, so TEAEs and MAAEs are the same.
